# A Complement Atlas identifies interleukin 6 dependent alternative pathway dysregulation as a key druggable feature of COVID-19

**DOI:** 10.1101/2023.03.25.23287712

**Authors:** Karel F.A. Van Damme, Levi Hoste, Jozefien Declercq, Elisabeth De Leeuw, Bastiaan Maes, Liesbet Martens, Roos Colman, Robin Browaeys, Cédric Bosteels, Stijn Verwaerde, Nicky Vermeulen, Sahine Lameire, Nincy Debeuf, Julie Deckers, Patrick Stordeur, Martin Guilliams, Sjoerd T.T. Schetters, Filomeen Haerynck, Simon J. Tavernier, Bart N. Lambrecht

## Abstract

To improve COVID-19 therapy, it is essential to understand the mechanisms driving critical illness. The complement system is an essential part of innate host defense that can also contribute to injury. All complement pathways have been implicated in COVID-19 pathogenesis, however the upstream drivers and downstream consequences on tissue injury remain ill-defined. Here, we demonstrate that complement activation is mediated by the alternative pathway and we provide a comprehensive atlas of the alterations in complement around the time of respiratory deterioration. Proteome and single-cell sequencing mapping across cell types and tissues reveals a division of labor between lung epithelial, stromal and myeloid cells in the production of complement, in addition to liver-derived factors. Upstream, IL-6 drives complement responses, linking complement dysregulation to approved COVID-19 therapies. In an exploratory proteomic study, C5 inhibition improves epithelial damage and markers of disease severity. Collectively, these results identify complement dysregulation as a key druggable feature of COVID-19.

## Introduction

Respiratory diseases have an enormous impact on human health. Numerous emerging pathogens exploit the lungs as a portal of entry to cause infection and enable transmission, as exemplified by the 2002 Severe Acute Respiratory Syndrome (SARS) outbreak, the 2012 Middle East Respiratory Syndrome (MERS) outbreak and the Coronavirus disease 2019 (COVID-19) pandemic, that all cause severe lung injury^1^. To improve therapies, it is essential to understand the mechanisms driving critical illness.

Initially, COVID-19 is characterized by a poor antiviral response, which is caused by both host and viral factors^2^. Uncontrolled viral replication leads to an excessive inflammatory response in the lungs, which impairs gas exchange and damages the microcirculation. Beyond supportive measures, treatment in the hyperinflammatory phase of COVID-19 consists of corticosteroids^3^, interleukin 6^4^ (IL- 6) and Janus kinase^5^ (JAK) inhibitors. Despite the benefit of these therapies, elderly and immunocompromised persons remain at risk for serious complications, even in the context of vaccination and less virulent variants^6^. Thus, more effective treatments could improve outcome and prevent long-term sequalae.

The complement cascade is an indispensable and multifunctional arm of the innate immune system^7, 8^. It contributes to pathogen recognition and phagocytotic clearance, recruits and activates immune cells, and induces direct cell lysis via the membrane attack complex (C5b-9, MAC)^7, 8^. Complement responses can be initiated via three pathways (Fig. 1a). The classical and lectin pathways are triggered by immune complexes and pathogen recognition molecules respectively, leading to cleavage of C4 and C2. The spontaneous hydrolysis of C3 triggers further activation of native C3 to set off the alternative pathway. All modes of complement activation converge on the proteolysis of C3 and C5, generating the potent pro-inflammatory peptides C3a and C5a, while C5b initiates MAC formation.

**Figure 1:**
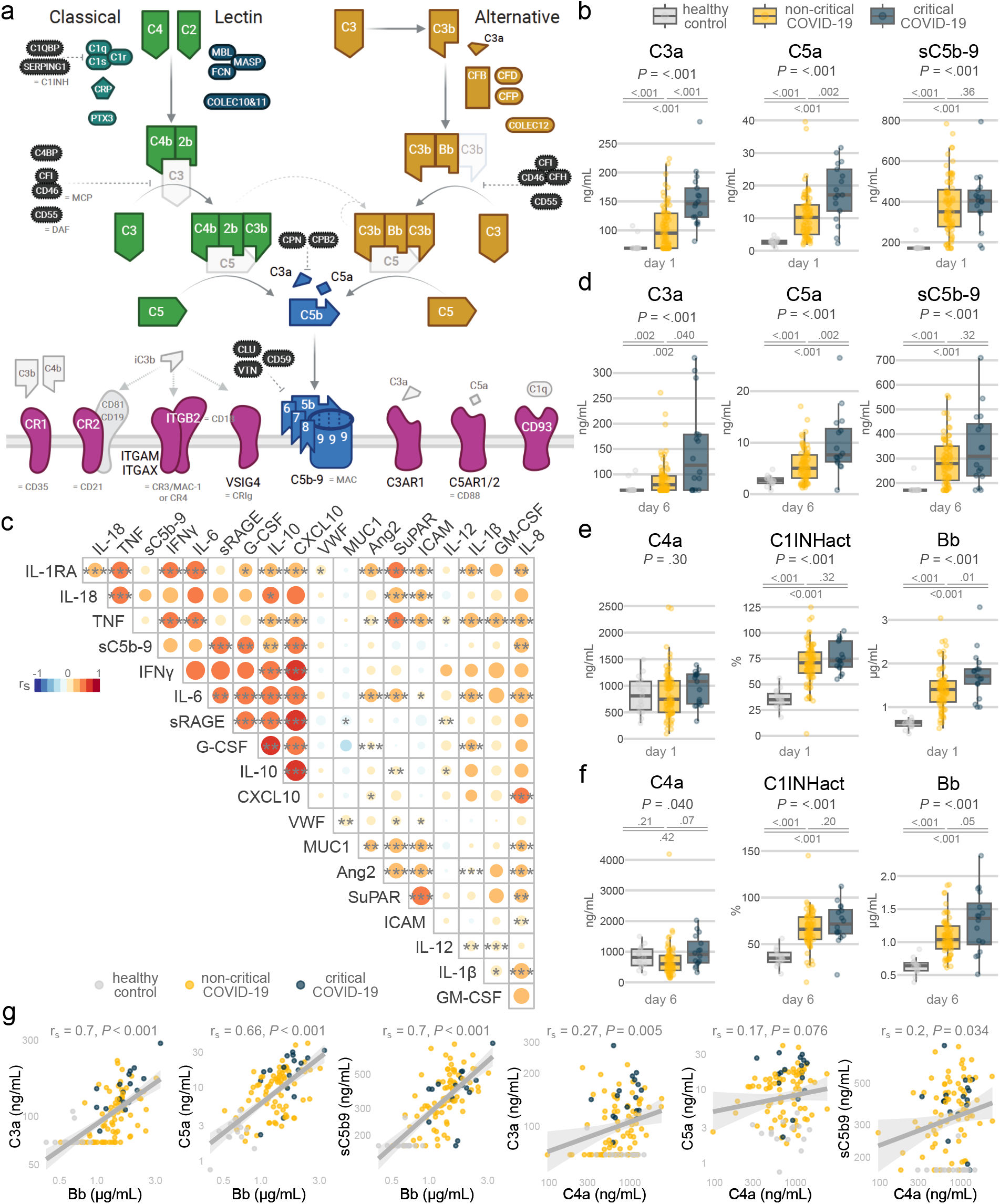
Complement activation via the alternative pathway correlates to disease severity. a, Schematic representation of the complement system. The classical and lectin pathways are colored in green, the alternative in yellow, the terminal in blue. The complement regulators are represented in black and the main complement receptors in purple. Created with BioRender.com. b, d, e, f, Plasma levels of activated complement components at day 1 (b, e) and day 6 (d, f) in COVID- 19 patients or age-matched healthy controls. Critical illness is defined as the need for invasive mechanical ventilation or death at any time during the trial follow-up. The P values are calculated using Kruskal-Wallis tests, with post-hoc pairwise Benjamini-Hochberg corrected Wilcoxon rank sum tests. C1INHact: functional activity of C1 inhibitor. Healthy, n = 15; day 1 non-critical, n = 76; day 1 critical, n = 17; day 6 non-critical, n = 72; day 6 critical, n = 16. c, Correlation matrix of hierarchically clustered biomarkers, measured in serum of COVID-19 patients (ZILUCOV trial participants at day 1, 6 and 15 or hospital discharge). Correlations are calculated by two-sided Spearman’s rank tests for pairwise complete observations, adjusted for multiple testing by the Benjamini-Hochberg procedure and indicated by *, P < .05; **, P < .01; ***, P < .001. The circle area and color represent the Spearman’s rank correlation coefficient. g, Linear correlations for Bb (left) and C4a (right) in relation to the terminal complement components C3a, C5a and soluble C5b-9. The data points are colored according to the disease severity. Correlation coefficient and P values are determined by a two-sided Spearman’s rank test.

To prevent inappropriate immune activation and collateral damage to tissues, complement responses are tightly controlled under homeostatic conditions. The key importance of complement and its regulation is best illustrated by genetic complement deficiencies^9, 10^, which lead to immunodeficiency, auto-immunity, endothelial damage and/or kidney injury. In addition to these inborn disorders, dysregulation of the complement system has been reported across a spectrum of infectious diseases, including those caused by emerging coronaviruses^11–16^.

All three complement arms were shown to be activated in COVID-19^17–24^, however their relative contributions remain ill-defined. While complement was initially described as a liver-derived cascade, recent studies have demonstrated unexpected cell- and tissue-specific production and functions for the complement system^8, 25–30^. Whether these novel roles for the complement components are also at play in COVID-19, is still poorly understood. A number of randomized trials explored the effects of targeting various components of the complement cascade^31–34^. Despite the encouraging results of some of these trials, a comprehensive analysis of the immunological consequences of the complement activation and its therapeutic targeting remain lacking.

In this study, we combined proteomics, single cell transcriptomics and immuno-assays to construct a COVID-19 Complement Atlas. We display the profound complement dysregulation, which correlates to disease severity and is best explained by alternative pathway activation. Using various single cell datasets, we map complement gene expression across tissues and cell types. Our results reveal a division of labor between locally sourced complement factors derived from pulmonary stromal, epithelial and myeloid cells, in addition to liver-derived factors. Comparing clinical samples before and after therapeutic IL-6 blockade, we identify IL-6 as a crucial upstream regulator of complement production and activity. Finally, we highlight the improved severity signature in the proteome of COVID-19 patients after treatment with zilucoplan, a C5 inhibitor. Taken together, our integrated COVID-19 Complement Atlas establishes how complement dysregulation contributes to the pathogenesis of COVID-19.

## Results

Complement activation is an intricate process resulting in the generation of cleaved C3, C5 and subsequent assembly of the MAC. To investigate whether complement dysregulation is involved in the pathogenesis of COVID-19, we measured activated complement components in the plasma of COVID-19 patients that participated in the COV-AID or SARPAC clinical trials^35, 36^ (Suppl. Table 1 and 2). Both multicenter clinical trials enrolled hospitalized COVID-19 patients with hypoxia in the course of 2020 (hypoxia was defined as a PaO_2_/FiO_2_ below 350; see Methods). Patients on mechanical ventilation or with high serum concentrations of inflammatory markers were not eligible for SARPAC; patients with high concentrations of inflammatory serum markers were included in COV-AID, providing they were not on mechanical ventilation longer than 24h at randomization. In this analysis, we found that the plasma concentration of C3a, C5a and soluble MAC (sC5b-9) was profoundly elevated in COVID-19 patients, compared to age-matched healthy controls (Fig. 1b). We also compared patients with or without a critical disease course, defined as the need for mechanical ventilation or death at any time. In comparison to non-critical patients, critical patients had additional increases of C3a and C5a, while this did not reach significance for sC5b-9. These results indicate ongoing complement activation at the time of respiratory deterioration in COVID-19, in line with previous reports^11, 16, 19, 21^.

We wondered if the increased complement activity would also correlate with established hallmarks of COVID-19^37^. Indeed, C-reactive protein (CRP), ferritin and D-dimer concentration at day 1 correlated with activation of the terminal complement pathway, especially C3a (Suppl. Fig. 1a). Higher C3a and sC5b-9 were correlated with worse oxygenation at trial inclusion (Suppl. Fig. 1b). We additionally measured biomarkers, including sC5b-9, during the disease course of the patients in the ZILUCOV clinical trial, which included hypoxic and hyperinflammatory patients with the same clinical profile as COV-AID, to evaluate anti-C5 therapy in COVID-19 (see Methods)^34^. sC5b-9 correlated with established biomarkers of disease severity^37–39^, such as the soluble receptor for advanced glycation end products (sRAGE), granulocyte-colony stimulating factor (G-CSF), interleukin 10 (IL-10), C-X-C motif chemokine ligand 10 (CXCL10) and interleukin 8 (IL-8 or CXCL8) (Fig. 1c). In fact, sC5b-9 hierarchically clustered with primary inflammatory cytokines such as tumor necrosis factor alpha (TNFα), interferon gamma (IFNγ) and interleukin 6 (IL-6). These correlated markers were clearly elevated in non-survivors versus survivors (Suppl. Fig. 1c), suggesting a central role for complement in inflammation induced by COVID-19.

Next, we investigated if complement activation persisted in the disease course. Regardless of treatment allocation in the SARPAC and COV-AID trials, levels of C3a and C5a remained higher in the critically ill participants at day 6, compared to the non-critically ill patients (Fig. 1d). These activated complement factors were also notably increased at day 6 in the patients who would eventually pass away (Suppl. Fig. 1d). While cleaved complement components correlated well with disease severity, the levels of native complement proteins did not predict disease severity (Suppl. Fig. 1e-f). Together, these data indicate that complement activation is a key sustained feature of COVID-19.

Three distinct pathways, defined as classical, lectin, and alternative pathways, can initiate the complement cascade. To investigate which of these is predominant in COVID-19, we measured upstream complement factors. Concentration of C4a was not raised in COVID-19 patients compared to age-matched healthy controls at both day 1 and day 6, suggesting little activity of the classical and lectin pathway (Fig. 1e-f). C1 inhibitor activity markedly increased in COVID-19, potentially reducing classical and lectin activation in COVID-19, and COVID-19 specific antibodies were also low in most patients at trial enrollment, despite already high complement activity (Suppl. Fig. 1g). In contrast, the Bb fragment of complement factor B (Bb) was greatly elevated in the plasma of participants of COV- AID and SARPAC at both time points, indicative of increased alternative pathway activity (Fig. 1e-f, Suppl. Fig. 1d). Similar to C3a and C5a, Bb was higher in critically ill patients than in non-critical patients at both time points. When terminal pathway activation was read out, either as the concentration of plasma C3a, C5a or sC5b9, cleaved factor B was a better predictor for activity than C4a (Fig. 1g), identifying alternative pathway activation as the main amplifier of complement in COVID-19.

The complement system is a proteolytic cascade, and its activity can be suppressed due to excessive complement factor consumption. To study functional complement activity, we selected serum samples of patients in three clinical trials which received glucocorticoid therapy^3^ (Suppl. Table 3). Compared to age-matched healthy controls, the complement activity of all three pathways did not decrease in COVID-19 patients at trial inclusion, and there was no correlation between the functional pathway activities (Suppl. Fig. 2a-b). However, functional alternative pathway was lower in non-surviving versus surviving COVID-19 patients (Fig. 2a-b), consistent with most alternative pathway consumption in the most critically ill patients.

**Figure 2:**
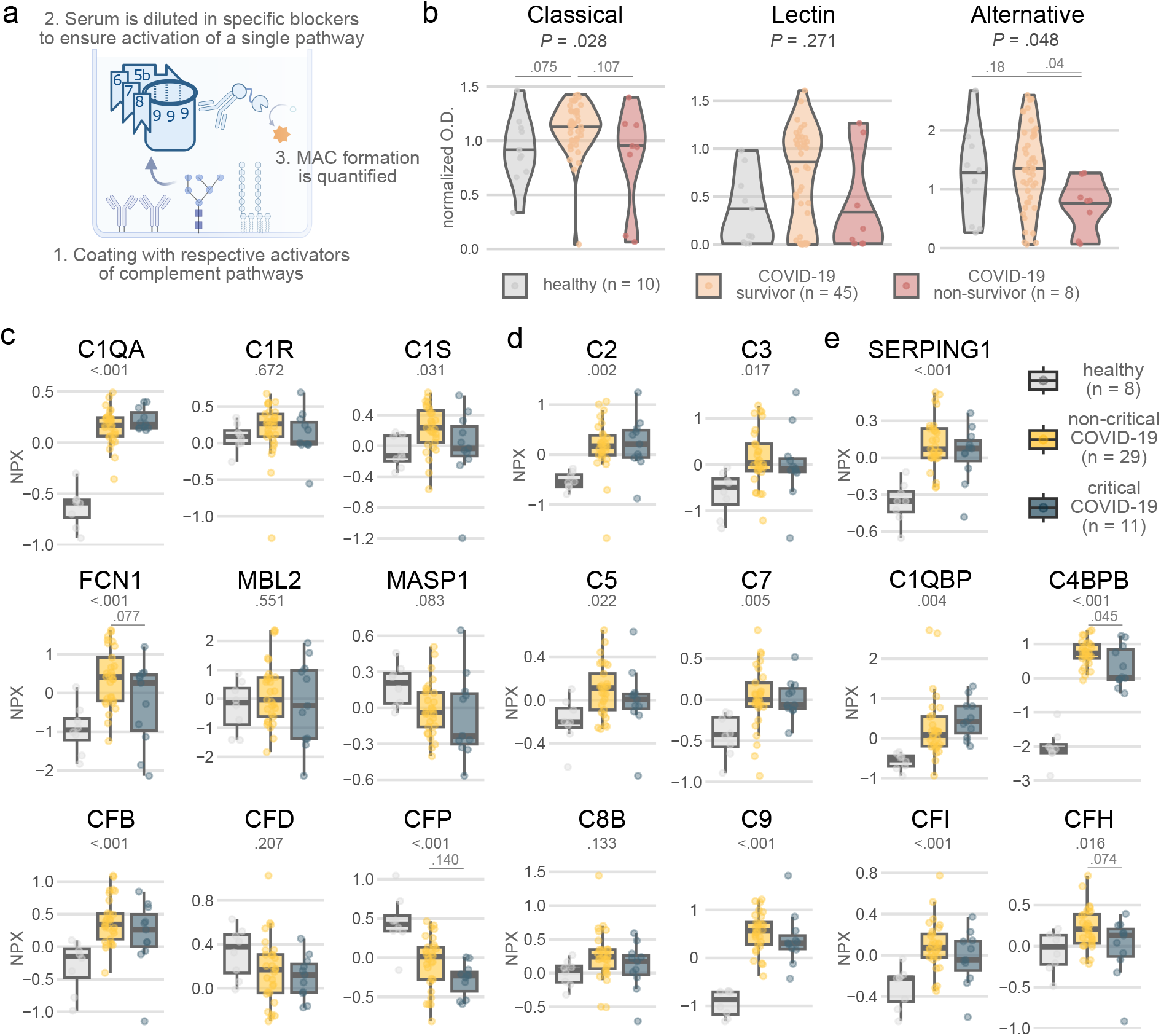
Complement activities and proteomics reveal consumption of alternative pathway factors in COVID-19. a, Schematic of the assay to quantify the activity of each complement pathway. Created with BioRender.com. b, Violin plots of the activity for each complement pathway in COVID-19 patients (at trial inclusion) or age-matched controls. P values are determined by Kruskal-Wallis tests and follow-up pairwise Benjamini-Hochberg-corrected Wilcoxon rank sum tests. Activity levels are measured as optical densities (O.D.) and normalized for the positive control. c, d, e, The upstream complement activators (c), main complement proteins (d) and complement regulatory proteins (e) in COVID-19 (ZILUCOV trial participants at the trial inclusion), measured by the proximity extension assay. P values are calculated by a one-way ANOVA with Tukey’s Honest Significant Difference method as post-hoc test. Protein levels in serum are plotted as Normalized Protein eXpression (NPX) on a log2 scale.

To thoroughly map complement proteins in serum of COVID-19 patients, we made use of the Olink Explore 3000 technology, a proximity extension assay with next-generation sequencing read- out of 2921 proteins on the serum of COVID-19 patients (for method validation and patient selection, see Methods, Suppl. Fig. 2c-d and Suppl. Table 4). At baseline, various complement proteins were upregulated in these participants of the ZILUCOV trial, including the complement cascade-initiating proteins C1QA, C1S, pentraxin 3 (PTX3), ficolin 1 (FCN1) and complement factor B (CFB) (Fig. 2c and Suppl. Fig. 2e). Complement factor properdin (CFP), which stabilizes the alternative pathway C3 and C5 convertases, was reduced in COVID-19 patients. This decrease likely corresponds with properdin consumption during complement activation, being the major rate-limiting complement factor in serum for the alternative pathway (Suppl. Fig. 2f), and its consumption during COVID-19. Similarly, C2 and mannose binding lectin 2 (MBL2) were the main determinants for the functional classical and lectin pathway activities (Suppl. Fig. 2f). The levels of total C2, C3, C5, C7 and C9 were higher in COVID-19 (Fig. 2d). With the exception of the complement receptor 1 (CR1), regulatory proteins such as C1 inhibitor, C1Q and C4 binding proteins (C1QBP, C4BPB) and complement factor I (CFI) were also elevated in COVID-19 (Fig. 2e and Suppl. Fig. 2g-h). For these measurements, which likely detect both native as well as activated proteins, we found no differences between patients with or without a critical disease course, which was in contrast to levels of activated complement components (Fig. 1a,d). Thus increased synthesis of complement factors seems to compensate for its consumption during COVID-19.

While serum complement is mostly considered liver-derived, lung and diverse other cell types have recently been described as important sources of complement^7, 27, 28, 40–42^. To identify putative local sources of complement in healthy and COVID patients, we broadly mapped complement gene expression. Since COVID-19 is characterized by respiratory failure, we sourced a single-nucleus RNA sequencing lung atlas which contains samples from COVID-19 autopsies as well as healthy lung biopsies^43^ (Fig. 3a-b, Suppl. Fig. 3a). We further focused on the alveolar space and queried broncho- alveolar lavage (BAL) cells in a CITE-sequencing experiment which our lab reported previously^35^ (Fig. 3c-d, Suppl. Fig. 3b). To study cells in circulation, we analyzed single-cell RNA sequencing of fresh blood^44^, which also includes neutrophils, often being absent from other datasets (Fig. 3e-f, Suppl. Fig. 3c). While the liver is an important source of complement, only one single-nucleus RNA sequencing dataset of COVID-19 liver samples has been published, which does not contain control samples^45^. To enable comparisons with non-infected patients, we merged this dataset with a liver atlas which we published in 2022^46^ (Fig. 3g, Suppl. Fig. 3d-e). Collectively, these datasets form a resource for complement expression across tissues and cell types at a single-cell resolution, pointing out that complement factors are widely generated beyond the liver. This production throughout the human body, likely allowing different tissues to respond according to their specific needs, highlights the evolutionary conservation of complement not only across species^8^, but also across organs.

**Figure 3:**
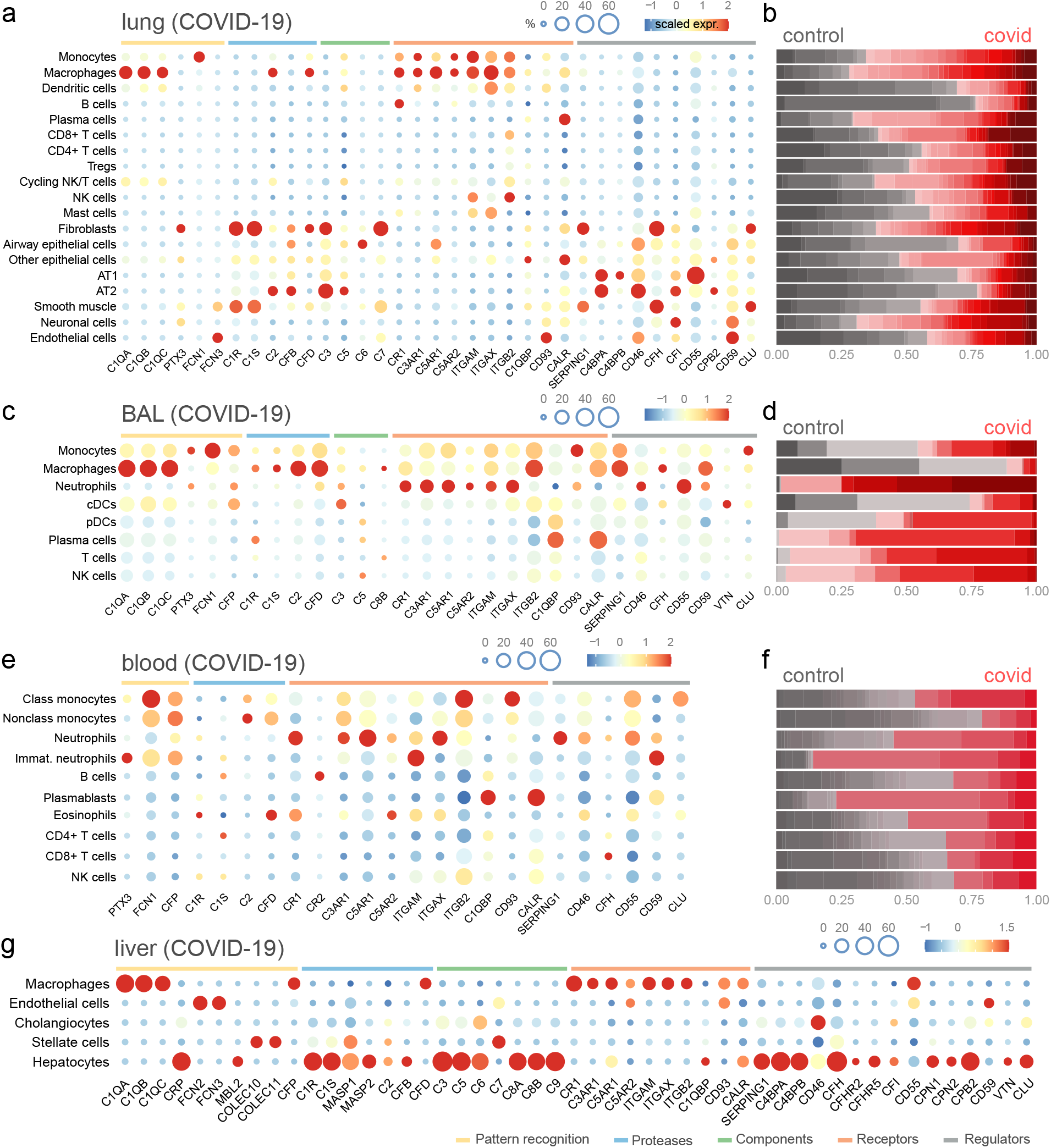
An atlas of complement gene expression across tissues and cell types in COVID-19. a, Complement gene expression across lung cell types in deceased COVID-19 patients, from the single-nucleus RNA sequencing lung atlas by Melms et al., 2021^43^. The dot size represents the proportion of cells expressing complement genes, while the color scale represents the average expression level. b, Relative contribution of control versus COVID-19 samples to the total cell numbers in lung (panel a). Each shade represents an individual patient, with control samples in grey and COVID-19 samples in red. c, Gene expression of complement factors in the broncho-alveolar lavage (BAL) of COVID-19 patients, from the a CITE-sequencing experiment which we have previously published^35^. d, Relative cell proportions of control versus COVID-19 samples for each cell type in BAL (panel c). e, Complement expression in fresh peripheral-blood mononuclear cells and whole blood of COVID-19 patients, in the single-cell RNA sequencing dataset by Schulte-Schrepping et al., 2020^44^. f, Relative contribution of control versus COVID-19 samples to the total number of cells in blood (panel e). g, Expression of complement genes across major liver cell types in COVID-19 patients. The data are derived from the Harmony-integrated^71^ dataset containing the single-nucleus RNA sequencing of lethal liver samples by Delorey et al., 2021^45^ and CITE- and single-nucleus sequencing data from an atlas containing both lean and obese liver biopsies^46^. Only single-nucleus sequencing data was retained to quantify complement expression per cell type.

This atlas enabled us to identify patterns in the cell-type specific production of complement components, and understand the driving forces behind the alternative pathway dysregulation in COVID-19. These datasets showed that for the alternative pathway, mainly fibroblasts and alveolar type 2 cells (AT2s) expressed C3, CFB and complement factor D (CFD) (Fig. 4a-c). Lung macrophages and neutrophils also produced CFD, while there was only little production of properdin (CFP) in the lung (Fig. 4c, Suppl. Fig. 4a-c). Instead, properdin was predominantly derived from myeloid cells in circulation (Fig. 4d-e). Similar to the alternative pathway, classical factors were expressed by various cell types in the lung. C1QA, C1QB and C1QC were produced by macrophages (Fig. 4f), whereas C1R and C1S was made by stromal cells in the liver and lung (Suppl. Fig. 4d), suggesting cross-lineage collaboration for the assembly of C1 complement complex. C2 mainly originated from macrophages and epithelial cells in the lung (Fig. 4f), while we curiously could not find C4A/B expression in any of the datasets (Fig. 3). Monocytes and endothelial cells in liver and lung produced lectin pathway-activating ficolins, while mannose-binding lectins (MBL) and MBL-associated serine proteases (MASP) predominantly stemmed from hepatocytes (Fig. 3g). For all three pathways, these data highlight the local generation of complement, through a division of labor between stromal, epithelial and myeloid cells.

**Figure 4:**
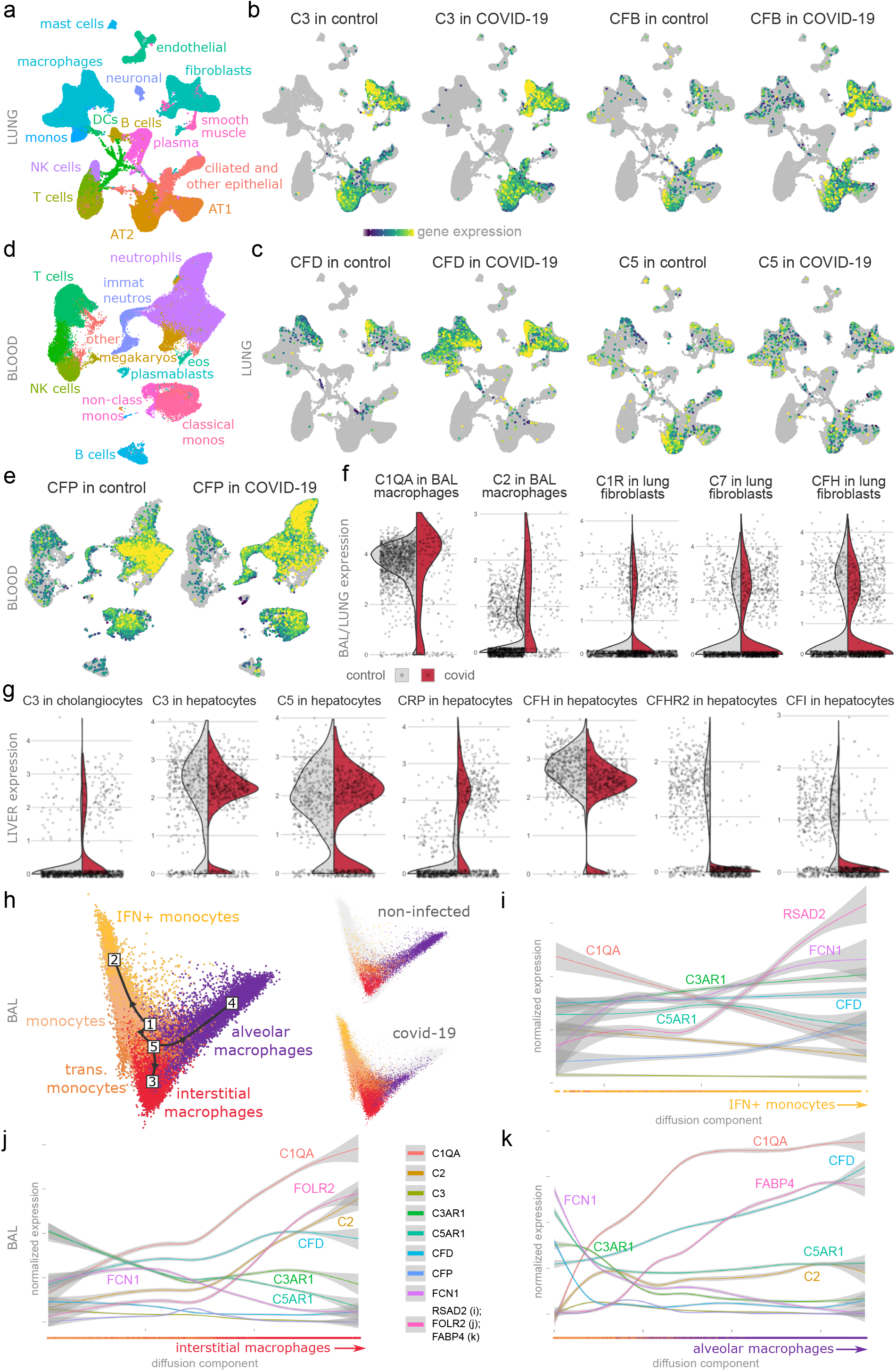
Complement production is divided between lung stroma, epithelium, myeloid cells and liver cells. a, Uniform manifold approximation and projection (UMAP) of the lung single-nucleus RNA sequencing experiment, colored by cell type annotation. b, c, Lung RNA expression of complement component 3 (C3) and complement factor B (CFB) (b), complement factor D (CFD) and C5 (c), separated by COVID-19 status. d, UMAP of blood single-cell RNA sequencing, and colored according to the cell type annotation. e, Expression of complement factor properdin (CFP) in blood, split for control or COVID-19 patients. f, g, Gene expression of complement factors in macrophages and fibroblasts in broncho-alveolar lavage (BAL) and lung (f) or in liver (g). Gene expression is represented as violin plots, split by COVID- 19 status. A proportional number of cells was plotted per condition and was overlaid as dots. h, Diffusion map and Slingshot trajectory analysis of monocytes and macrophages in BAL. Monocytes differentiate in either interferon-stimulated (IFN+) monocytes (1 to 2), or via transitional (trans.) monocytes into interstitial (1 to 5 to 3) and alveolar macrophages (1 to 5 to 4). The close-ups feature the diffusion maps of either non-infected or COVID-19 patients. i, j, k, Expression of complement genes across the diffusion components for the trajectories towards interferon-stimulated monocytes (i), interstitial macrophages (j) or alveolar macrophages (k).

For the terminal factors, we observed little C5 expression in the lungs (Fig. 4c, Suppl. Fig. 4c), while there was strongly expression in the liver (Fig. 4g), suggesting mostly extrapulmonary sources for C5. Locally, C6 was produced by ciliated cells, (Suppl. Fig. 4e), and C7 by fibroblasts and smooth muscle cells (Fig. 4f and Suppl. Fig. 4d). Hepatocytes produced C6, C8 and C9, while liver stellate cells expressed C7 (Fig. 3). In terms of regulators of the alternative and terminal pathways, CFI was locally expressed by fibroblasts, neuronal cells and epithelium. Complement factor H (CFH) was expressed by fibroblasts and smooth muscle cells, complement receptor 1 (CR1, CD35) by myeloid or B cells and membrane cofactor protein (MCP, CD46) by multiple cell types. Decay-accelerating factor (DAF, CD55) and MAC inhibitory protein (CD59) were also produced by various cell types. Surprisingly, C5AR1 was expressed on ciliated cells, in addition to its well-known expression on neutrophils and macrophages (Suppl. Fig. 2f-h). In general, the distribution of complement genes was similar in COVID-19 and non-infected controls. Only for complement factor H-related protein 2 (CFHR2) and CFI, there was decreased hepatic expression in COVID-19 patients (Fig. 4g), although these proteins were not reduced in serum of hospitalized COVID-19 patients (Fig. 2e, Suppl. Fig. 2h). This suggests that the observed increases in serum complement proteins are not caused by changes in RNA expression, but could be due to post-translational changes and/or increased numbers of complement producing cells upon SARS-CoV-2 infection. Differences in timing between blood collection (around time of respiratory deterioration) and liver and lung sampling (post-mortem) might also contribute to these findings.

COVID-19 is characterized by profound changes in myeloid compartment with a loss of alveolar macrophages^35, 47–49^. This is accompanied by a massive influx of complement receptor-expressing myeloid cells^14, 35, 49^ into the alveolar space which upregulated complement gene signature (Suppl. Fig. 4h-i). To study how these incoming myeloid cells participate in complement production, we used a previously published^35^ Slingshot trajectory analysis^50^ (Fig. 4h). In this analysis, FCN1^+^ monocytes differentiated in RSAD2^+^ interferon-stimulated (IFN^+^) monocytes (trajectory 1 to 2), via CHI3L1^+^ transitional monocytes into FOLR2^+^ interstitial macrophages (IMs, trajectory 1 to 5 to 3) or into FABP4^+^ alveolar macrophages (AMs, trajectory 1 to 5 to 4)^35, 51^. Within these trajectories, expression of FCN1 and the anaphylatoxin receptors C3AR1 and C5AR1 decreased in IMs and AMs, while this did not occur in IFN^+^ monocytes^35^ (Fig. 4i-k). Both IMs and AMs produced C1q components, and the increased C1q serum levels might reflect ongoing macrophages proliferation in COVID-19^51, 52^. Within macrophages, we observed a further specialization in the production of complement factors, as C2 was mainly expressed by IMs and CFD by AMs. Again, these data emphasize the specialization in the production of complement, even within cell types of a single differentiated lineage.

We next studied mechanisms behind the increase in complement in COVID-19 (Fig. 2 and Suppl. Fig. 2). For this purpose, we searched for upstream ligands linked to complement genes as downstream targets in the NicheNet database^53^. This yielded predicted ligands which could drive complement expression across tissues (Fig. 5a, Suppl. Fig. 5a). We then plotted the protein levels of these putative ligands in healthy controls and COVID-19 patients (Fig. 5b). From this analysis, IFNγ, IL- 6 and IL-10 might affect complement expression and also correlated both with COVID-19 status and severity. To further investigate whether any of these cytokines could control clinically relevant complement activation, we checked for correlations between activated complement components and cytokines at day 1 of COV-AID and SARPAC. While IFNγ and IL-10 correlated better with C5a, IL-6 was the only cytokine to correlate with all activated terminal complement components (Fig. 5c and Suppl. Fig. 5c-d). Consistent with the large and persistent increase in serum IL-6 in COVID-19 (Suppl. Fig. 1c, Fig. 5b), we found that various lung cell types involved in complement production exhibited an upregulated gene signature reflective of IL-6 activity, which can signal both via the IL-6 receptor (IL6R) or the IL-6 cytokine family signal transducer (IL6ST) (Fig. 5d-e). These cells included dendritic cells, fibroblasts and to a lesser degree macrophages and smooth muscle cells. These results suggest a role for sustained IL-6 signalling affecting complement dysregulation in COVID-19 patients with critical disease outcomes.

**Figure 5:**
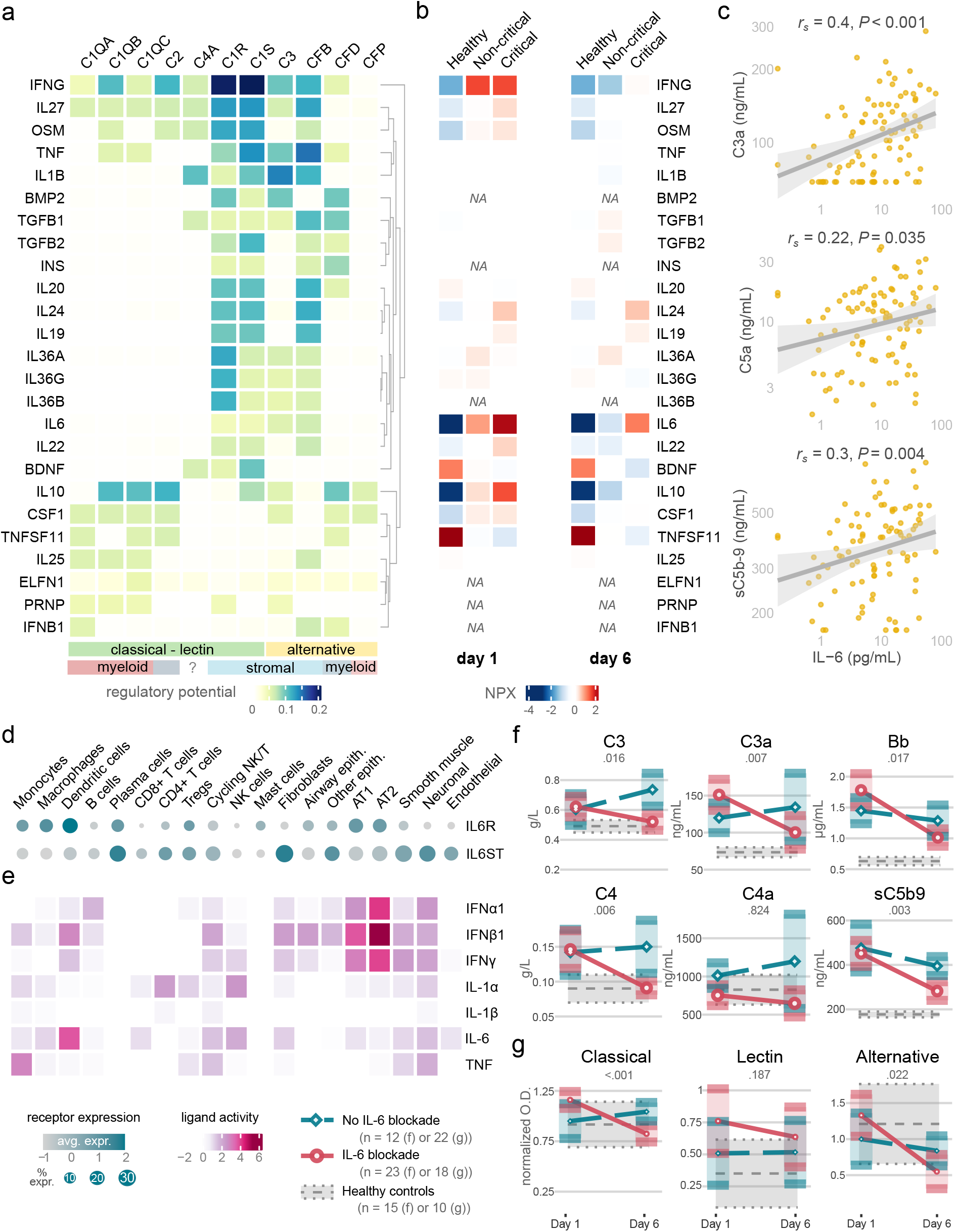
Interleukin 6 coordinates complement in COVID-19. a, The 25 ligands most likely to affect complement gene expression, with each ligand representing a row, each complement gene a column, and coloring according to the regulatory potential as predicted by the NicheNet ligand-target matrix^53^. b, Average levels in serum of putative ligands at day 1 (left) or day 6 (right) in healthy controls and COVID-19 patients, further divided according to disease severity. Protein levels are measured by proximity extension assay and expressed as Normalized Protein eXpression (NPX). NA: not available. c, Correlations of interleukin 6 (IL-6) in relation to C3a, C5a and sC5b-9. The correlation coefficients and P values are determined by two-sided Spearman’s rank tests. d, Expression of interleukin 6 receptor (IL6R) and interleukin 6 cytokine family signal transducer (IL6ST) across cell types in lung autopsies of COVID-19 patients. The dot size represents the fraction of cells expressing IL6R or IL6ST and the color represents the average scaled gene expression. e, Upregulated ligand activities of interferons, interleukin 1 (IL-1α/β), IL-6 and tumor necrosis factor (TNF) per cell type in the lungs of COVID-19 compared to control samples. f, The evolution of complement components in COVID-19 patients receiving IL-6 blockade (single dose of tocilizumab or siltuximab) or those who did not, between day 1 (predose) and 6. The data are plotted as averages with their 95% confidence interval. Both arms contain participants who received IL-1 blockade, due to the factorial design of COV-AID. g, Complement activity over time for each pathway, shown as averages with 95% confidence interval. Pathway activity is plotted as optical density (O.D.), normalized for the assay positive control. Control patients are COVID-19 patients from the ZILUCOV trial control arm, while the anti-IL-6 arm contains concurrent COV-AID participants who received anti-IL-6 therapy only (tocilizumab or siltuximab), without IL-1 blockade. P values for panels f and g are determined by a Wilcoxon rank sum test on the change between day 1 and 6.

To find a causal link between increased IL-6 and the complement system, we investigated whether anti-IL-6 drugs affected complement concentration and activity. Participants of the COV-AID trial who received either anti-IL-6 receptor (tocilizumab) or anti-IL-6 (siltuximab) had significantly lower levels of Bb, C3, C3a, C4 and sC5b-9 at 5 days after IL-6 blockade, compared to patients who did not receive anti-IL-6 drugs (Fig. 5f, Suppl. Fig. 5g). In contrast, daily recombinant interleukin 1 receptor antagonist (anakinra) did not affect complement levels in the same cohort of COV-AID patients, in contrast (Suppl. Fig. 5h-i). Due to the factorial design of the COV-AID trial^36^, both the anti-IL-6 and anti-IL-1 comparisons contain patients with the other intervention. To mitigate a potential interaction between both interventions, we validated these findings in a distinct cohort comprised of control patients from ZILUCOV and anti-IL-6(R) treated patients from COV-AID who did not receive anakinra (Suppl. Table 3). In accordance, classical and alternative pathway activity decreased significantly following anti-IL-6(R) therapy, while the level of functional lectin pathway did not drop significantly (Fig. 5f). Together, these data identify IL-6 as a key druggable regulator of complement responses in COVID-19.

Finally, we aimed to understanding the consequences of complement inhibition in COVID-19. In the ZILUCOV trial, daily administration of the C5 inhibitor zilucoplan did not significantly improve oxygenation at day 6 or 15, the primary endpoint (p = 0.12)^34^. However, a Bayesian analysis indicated that participants treated with C5 inhibition had an > 89% chance of an improved oxygenation compared to untreated patients, and an > 91% chance of better survival^34^. In addition, complement inhibition lowered serum IL-8 levels^34^, a granulocyte-attracting chemokine, which was also noted in an independent cohort treated with a C5a blocking antibody^54^. Despite these observations, the scope of biological responses affected by complement blockade in COVID-19 remains unexplored. For this purpose, we studied the evolution of the serum proteome between day 1 and 6 in a cohort of matched anti-C5 treated and untreated patients from the ZILUCOV trial, using the Olink Explore 3000 platform (see Methods, Suppl. Fig. 2d, Suppl. Table 4). No adjustments for multiplicity in the comparison between the anti-C5 and untreated arms were made, due to the exploratory nature of the analyses.

We first examined how the serum proteome differed at trial inclusion between patients with a critical and a non-critical disease course. Patients who required invasive mechanical ventilation or died during the trial had elevated levels of inflammatory mediators and of proteins typically expressed by epithelial cells (Fig. 6a), expanding on previous reports^38, 55, 56^. Next, when comparing the evolution between day 1 and 6 in control versus anti-C5 treated COVID-19 patients, several proteins changed differentially (Fig. 6b, Suppl. Fig. 6a) while both arms displayed decreases in pro-inflammatory and anti-viral signaling over time (Suppl. Fig. 6b). Many of the proteins with a differential evolution between both trial arms strongly correlated with critical illness (Fig. 6c), suggesting that complement inhibition could affect disease processes which correlate with COVID-19 severity. We validated the association of these proteins with disease severity in a separate cohort, which made use of the Olink 1500 platform^56^ (Suppl. Fig. 6c-d). In a paired analysis, we also observed a strong enrichment of severity-associated proteins which decreased significantly under the anti-C5 therapy but not in the control arm (Fig. 6d). Consistent with the profound decrease in sC5b-9 upon zilucoplan administration^34^, terminal complement components C7 and C8B levels in serum increased due to decreased consumption (Fig. 6e-f), but upstream complement activators were not affected (Suppl. Fig. 6e). Additional proteins which decreased under C5 blockade but not in untreated patients included markers of epithelial damage (AGR2, CAPS, SFTPA2) and inflammatory mediators (IL-6, IL-8, IL-15) (Fig. 6e-f), which have been used in previous trials as biomarkers of severe acute respiratory distress syndrome (ARDS)^57, 58^. Numerous nucleotide-binding proteins which are normally found only at low levels in serum (FUS, GADD45GIP1, JUN, SSB, ZHX2) declined more in anti-C5 treated patients (Fig. 6e-f), suggesting decreased spill of intracellular proteins into the extracellular space in a complement-dependent manner. Pathway analysis also pointed towards decreased damage and a reduced pro-inflammatory signaling upon treatment with anti-C5 therapy (Fig. 6g, Suppl. Fig. 6f). Together, these exploratory data suggest that complement C5 blockade around the time of respiratory deterioration reduce epithelial damage in COVID-19.

**Figure 6:**
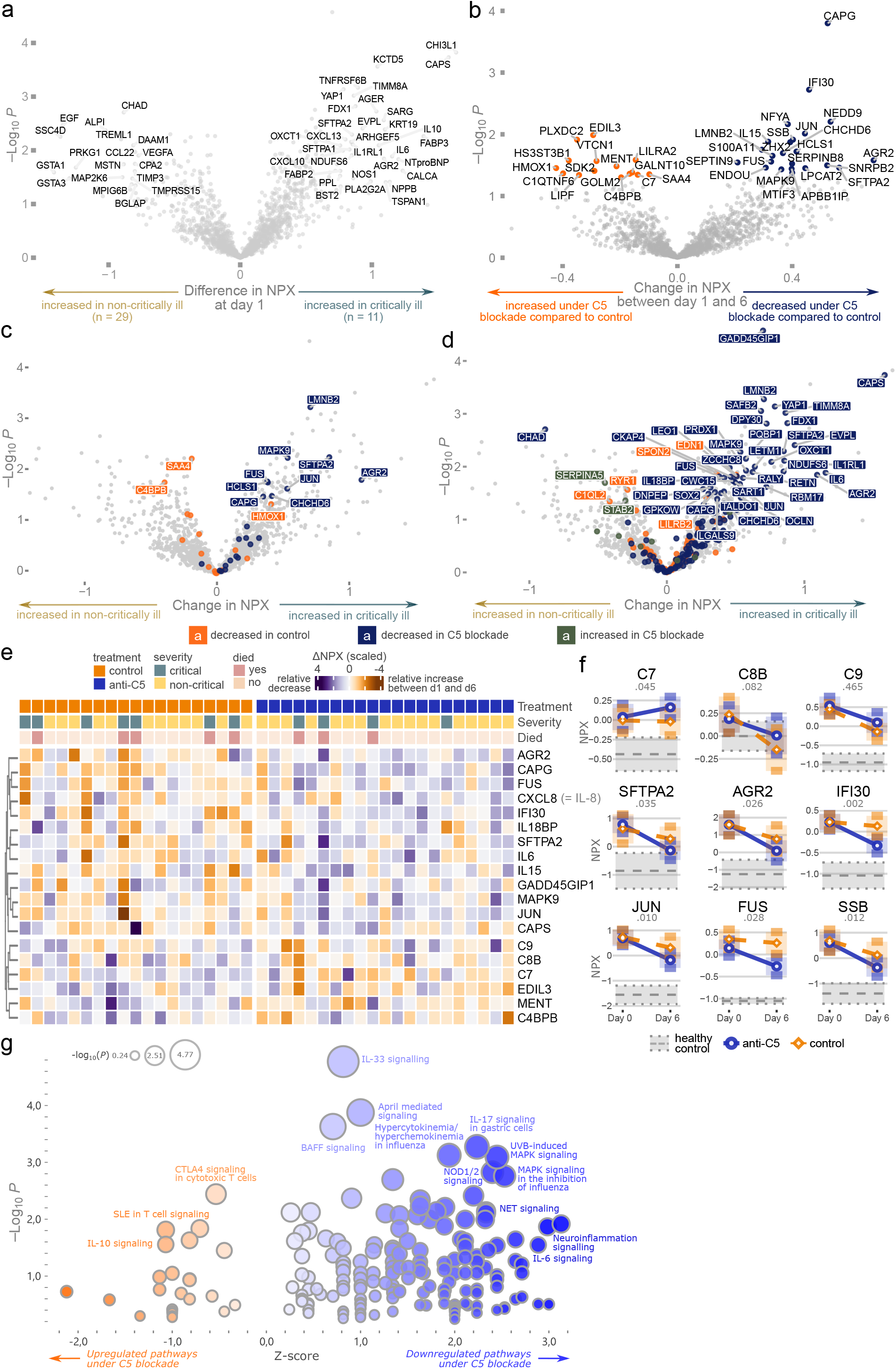
Complement inhibition improves the COVID-19 proteomic severity signature. a, Proteins associated with critical illness at day 1, defined as the need for invasive mechanical ventilation or death at any time during the trial. Proteins on the right are increased in patients with a critical COVID-19 disease course, while those on the left are decreased. The P values reflect two- sided t-tests without adjustment for multiplicity. The data are expressed as the difference in Normalized Protein eXpression (NPX) (which equals to a log_2_ fold change). b, Differences in the evolution of serum proteins between day 1 (predose) and day 6 of control versus anti-C5 treated patients. Proteins in orange display a relative increase more in anti-C5 treated patients compared to untreated patients, while proteins in blue have relative decrease under C5 inhibition. P values are determined by uncorrected two-sided t-tests. c, Proteins affected by anti-C5 therapy (panel b) projected on the critical illness-associated volcano plot (panel a). Orange proteins are relatively increased under C5 inhibition compared to untreated patients, while blue proteins are downregulated in anti-C5 treated patients, relative to untreated patients. d, Paired analysis for anti-C5 and control arm, plotted on volcano plot with critically illness-associated proteins. Proteins decreasing between day 1 and 6 in the anti-C5 arm only are labeled in blue, while those decreasing over time in the control arm only are plotted in orange. Two proteins increase only under anti-C5 (green) and no proteins increases in the control arm only. Two-sided t-tests with Benjamini-Hochberg adjustment are used to compare day 1 with day 6 protein levels for the untreated and the anti-C5 treated arm. e, Heatmap with hierarchically clustered proteins per patient, colored for the change in NPX between day 1 and 6. f, Changes of terminal complement proteins (top), severity-associated markers (middle) and nucleotide binding proteins (bottom) over time between anti-C5-treated or untreated patients. P values are calculated for the difference in log_2_ fold change over time with a two-tailed student t-test. g, Ingenuity Pathway Analysis of anti-C5 versus control arm. Pathways upregulated between day 1 and 6 under C5 blockade have a negative Z score (left, colored in orange), while those downregulated upon anti-C5 therapy have a positive Z score. P values are calculated with Fischer’s exact testing.

## Discussion

Despite vaccination, less virulent variants and evidence-based treatments, COVID-19 continues to be an important cause of disability, especially for people with weakened immune responses^6^. Thus, there remains an important need for interventions which benefit hospitalized patients. In addition, treatments are also urgently needed for other forms of acute respiratory distress syndromes, for which no disease-modifying therapies currently exist^59^.

Early on in the pandemic, dysregulated complement responses were linked to COVID-19^12–15^, and several randomized clinical trials were initiated^31–34^. Similarly, complement overactivation has been implicated in other forms of acute lung injury and sepsis as well^52, 60, 61^. Despite some promising clinical results in COVID-19, especially for C5a blockade^31^ and to a lesser degree for C5 blockade^34^, many questions remain on complement dysregulation in virus-induced acute lung injury. We show how excessive alternative pathway activation around the time of respiratory deterioration is coordinated by IL-6 signalling. This complement activation requires a complex collaboration between liver-derived and locally produced factors. Dysregulated complement responses contribute to epithelial damage and this seems to be improved upon C5 blockade, highlighting the therapeutic potential of complement inhibition.

To prevent inflammation and tissue damage in steady state, the complement cascade is tightly controlled by numerous proteins which limit its activation. In addition to these complement regulators, our results suggest an additional mechanism of checks and balances for its activation. While many complement factors are produced in the liver, we found that diverse cell types across tissues contribute to the generation of complement. This task separation between liver, myeloid, stromal and epithelial cells could serve as a brake on excessive complement production and subsequent activation by a single player. Given the importance of complement in host protection, as illustrated by various complement deficiencies^9, 10^, it also guarantees continuous complement production even in case of cell-specific disturbances. This specialization within the lung mirrors the zonation of the liver^46^, which influences hepatic complement production^62^. In combination with the 2921-plex proteomics, these data form an atlas of complement in healthy controls and COVID-19 patients, which might inform future studies.

In COVID-19, the complement system plays opposing roles. Early in the infection, adequate innate immune responses, including complement activation, contribute to a rapid return to homeostasis. To circumvent timely immune recognition, the SARS-CoV-2 virus employs multiple mechanisms, which include the disruption of epigenetic regulation^63^ and inhibition of complement^64^ by its ORF8 protein. Later in the disease course, defective viral clearance induces excessive lung inflammation, which is promoted by complement activation^65^. We found the alternative pathway to be particularly overactive in COVID-19 patients around the onset of respiratory failure, with consumption of CFP and large increases in Bb levels, predicting the levels of activated terminal components. At this time, the initial triggers of alternative pathway activation in COVID-19 still remain unclear. Its activation could be due to excess C3b generation in the presence of viral components^66^ or result from overwhelmed complement regulatory mechanisms in the context of widespread tissue damage. Although classical and lectin pathway activation can be perpetuated via the alternative pathway, initial triggering by immune complexes^18, 22^ or direct lectin binding^17, 24^ seems less likely given the unaltered levels of C4a around the time of respiratory failure.

Activation of the alternative pathway can amplify inflammation and tissue injury through multiple mechanisms. C3a and C5a promote key features of COVID-19, including myeloid cell recruitment and activation^42^, cytokine production^7^, T cell cytotoxicity^25, 26^, neutrophil extracellular trap formation and coagulopathy^13, 54^. Moreover, complement-driven myeloid migration into the alveolar space might induce a feed forward loop via myeloid derived IL-8, a chemokine which decreases upon inhibition of both C5^34^ and C5a^54^. Surprisingly, C5a might signal to ciliated cells and hepatocytes as well, which express C5AR1. Beyond anaphylatoxins, membrane attack complex formation can contribute to tissue damage through cell lysis as well as neutrophil recruitment^67^. The effectivity of C5a inhibition in COVID-19^31^ suggests that lung injury is primarily mediated by C5a however, with might co-signal via C5AR2 given the ineffectivity of C5AR1 blockade^33^.

Consistent with in silico predictions and transcriptomic signatures, we establish IL-6 as a crucial mediator of complement production and activity in COVID-19. These results are consistent with the established role of IL-6 in the coordination of acute-phase responses^68^, which is known to induce increases in complement levels. This observation also links the complement system to other approved therapies for COVID-19, such as JAK^5^ and IL-6 inhibitors^4^, suggesting that these therapies might alleviate tissue damage in part via their effects on the IL-6-complement axis. Compared to C5 inhibition, anti-IL-6 has a much slower impact on complement activity though^34^, and combining these therapies might have synergistic effects. Complement blockade might also ameliorate other forms of lung injury and sepsis, given available (pre)clinical evidence^61, 69^ and large increases of IL-6 in non-COVID-19 pneumonia and sepsis^70^. Taken together, these results point to complement activation as a key druggable feature of COVID-19.

While this study expands our understanding of up- and downstream processes involving alternative pathway dysregulation system in COVID-19, it also has limitations. All samples were collected during 2020, prior to vaccination and novel SARS-CoV-2 variants, which might alter complement responses. The small sample size and large number of proteins measured in the proximity extension assay led us to report P values uncorrected for multiple testing in comparison between anti-C5 and control patients. Thus, this analysis should be regarded as exploratory, hypothesis generating and requiring independent confirmation. Finally, our proteomic analysis on serum instead of plasma does not provide insight in the coagulation cascade, which is closely intertwined with complement and dysregulated during COVID-19 as well^13^. Ideally, future proteomic studies in ARDS should incorporate broncho-alveolar lavage and/or lung samples, since tissue-specific disturbances could be overlooked in serum or plasma. We were not able to analyze patient samples from non-COVID-19 respiratory failure or sepsis, but given the lack of disease-modifying therapies, it is of importance to investigate if the IL-6-complement axis also mediates lung injury in these forms of ARDS.

## Data Availability

All data produced in the present study are available upon reasonable request to the authors.

---

**Supplementary Figure 1:**
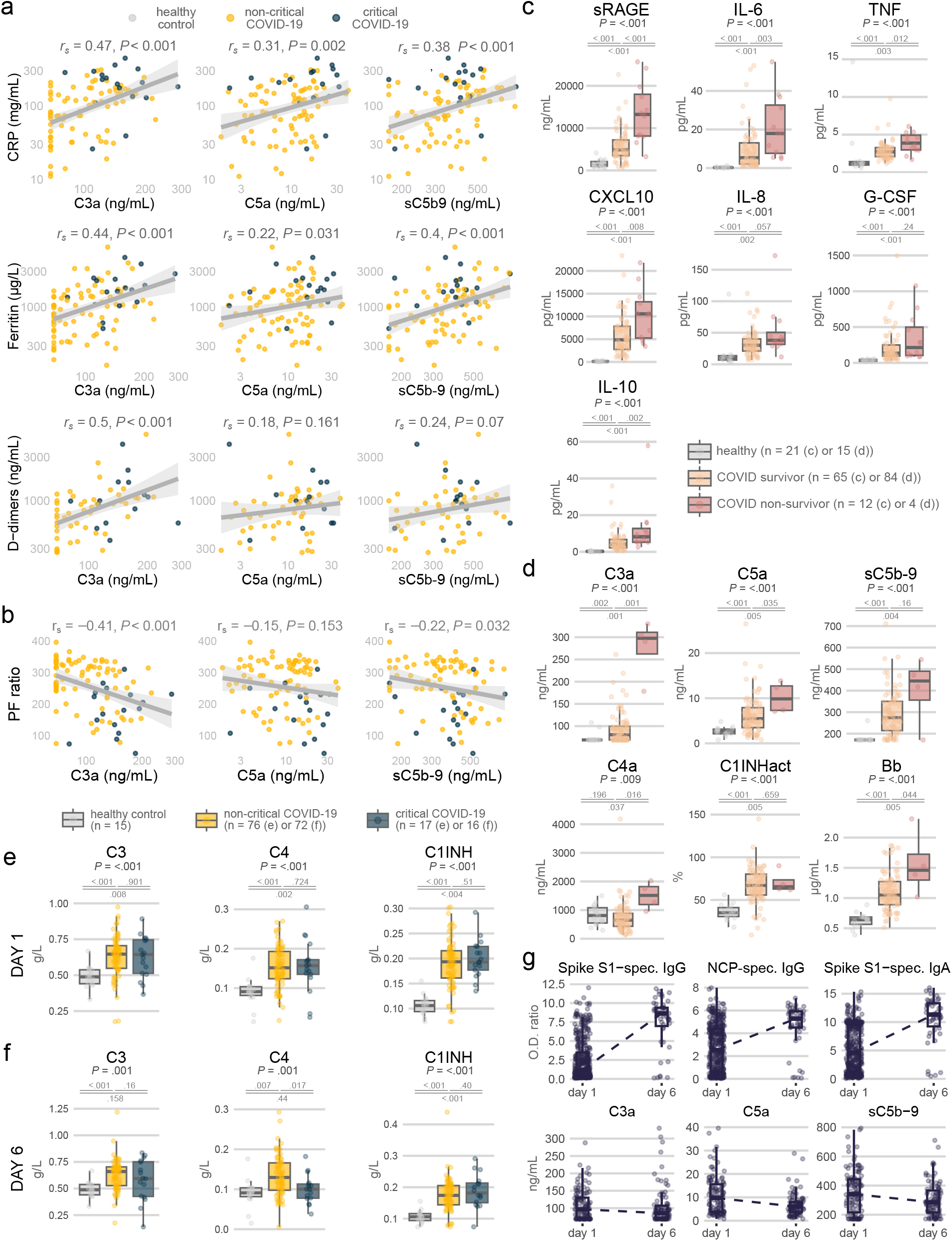
Complement activation via the alternative pathway correlates to disease severity. a, b, Linear correlations for the terminal complement components C3a, C5a and soluble MAC in relation to C-reactive protein (CRP), ferritin, D-dimers (a) and the partial pressure of arterial oxygen divided by the fraction of inspired oxygen (PF ratio) (b) at trial inclusion in COVID-19 patients (samples at inclusion of the SARPAC and COV-AID trials). Observations are colored by severity classification. Correlation coefficient and p-values are determined by a two-sided Spearman’s rank test. c, Serum levels of soluble receptor for advanced glycation end products (sRAGE), interleukin 6 (IL-6), tumor necrosis factor (TNF), C-X-C motif chemokine ligand 10 (CXCL10), interleukin 8 (IL-8 or CXCL8), granulocyte colony-stimulating factor (G-CSF) and interleukin 10 (IL-10) in healthy controls or COVID- 19 patients (ZILUCOV participants at trial inclusion), further subdivided as COVID-19 survivors and non-survivors. d, Levels of activated complement components at day 6 in COVID-19 survivors and non-survivors, in addition to healthy controls. C1INHact: functional activity of C1 inhibitor. e, f, Plasma levels of C3, C4 and C1 inhibitor at day 1 (e) and 6 (f). P values in panels c-f are determined by Kruskal-Wallis tests, with post-hoc pairwise Benjamini-Hochberg corrected Wilcoxon tests. g, Evolution of SARS-CoV-2 specific antibodies over time in comparison to the evolution of complement factors in plasma. NCP: Nucleocapsid.

**Supplementary Figure 2:**
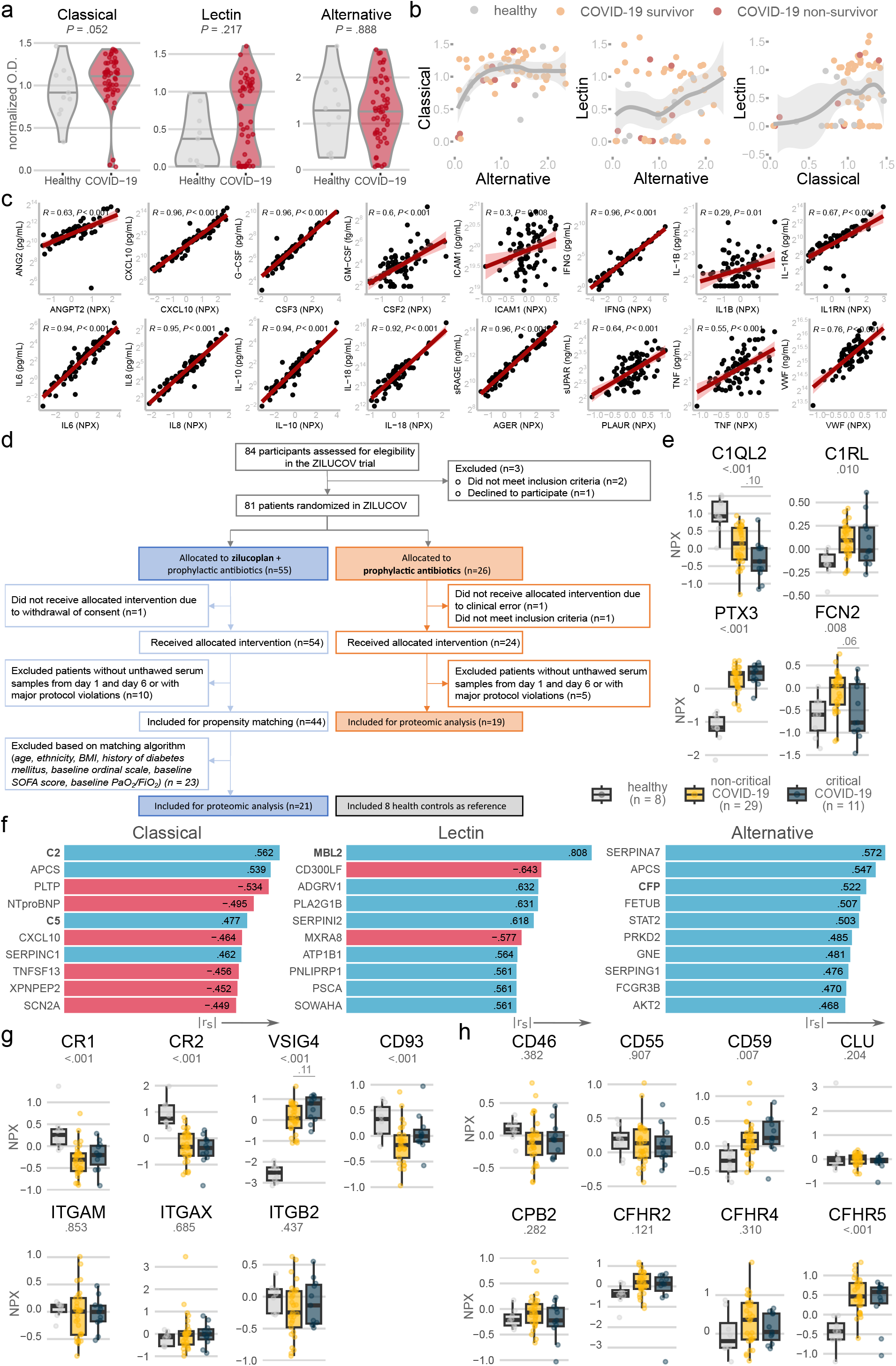
Complement activities and proteomics reveal consumption of alternative pathway factors in COVID-19. a, Violin plots displaying the complement activity for each pathway, for COVID-19 patients (at trial day 1) or age-matched controls. P values are determined by a two-sided test. Complement activities are quantified as optical densities (O.D.) and normalized for the positive assay control. b, Correlations between the functional complement pathway activities, with coloring of the data points according to the clinical outcome. A local regression fitting is overlaid with the scatterplot. c, Validation of the proximity extension assay, depicting correlations between protein levels measured by the proximity extension assay on the x-axis, and classic immune-assays on the y-axis. The correlation and P values are determined by two-sided Spearman’s rank tests. d, CONSORT flow diagram showing subject enrollment, allocation and inclusion in the proteomic study. e, g, h, Serum levels of upstream complement factors (e), complement receptors (g) and regulatory proteins (h) in COVID-19 patients (ZILUCOV trial participants at the trial inclusion), further separated according to disease severity. P values levels are calculated by one-way ANOVA tests with Tukey’s Honest Significant Difference method as post-hoc test. Protein levels in serum are plotted as Normalized Protein eXpression (NPX), which on a log2 scale. f, Best correlations between the complement pathway activities and proteins in serum, as determined by the proximity extension assay. Both day 1 and 6 samples of COVID-19 patients (ZILUCOV participants in the control arm) were included, as well as healthy controls. The bars are colored by the direction of Spearman’s rank correlation coefficient (blue: positive correlation; red: inverse correlation).

**Supplementary Figure 3:**
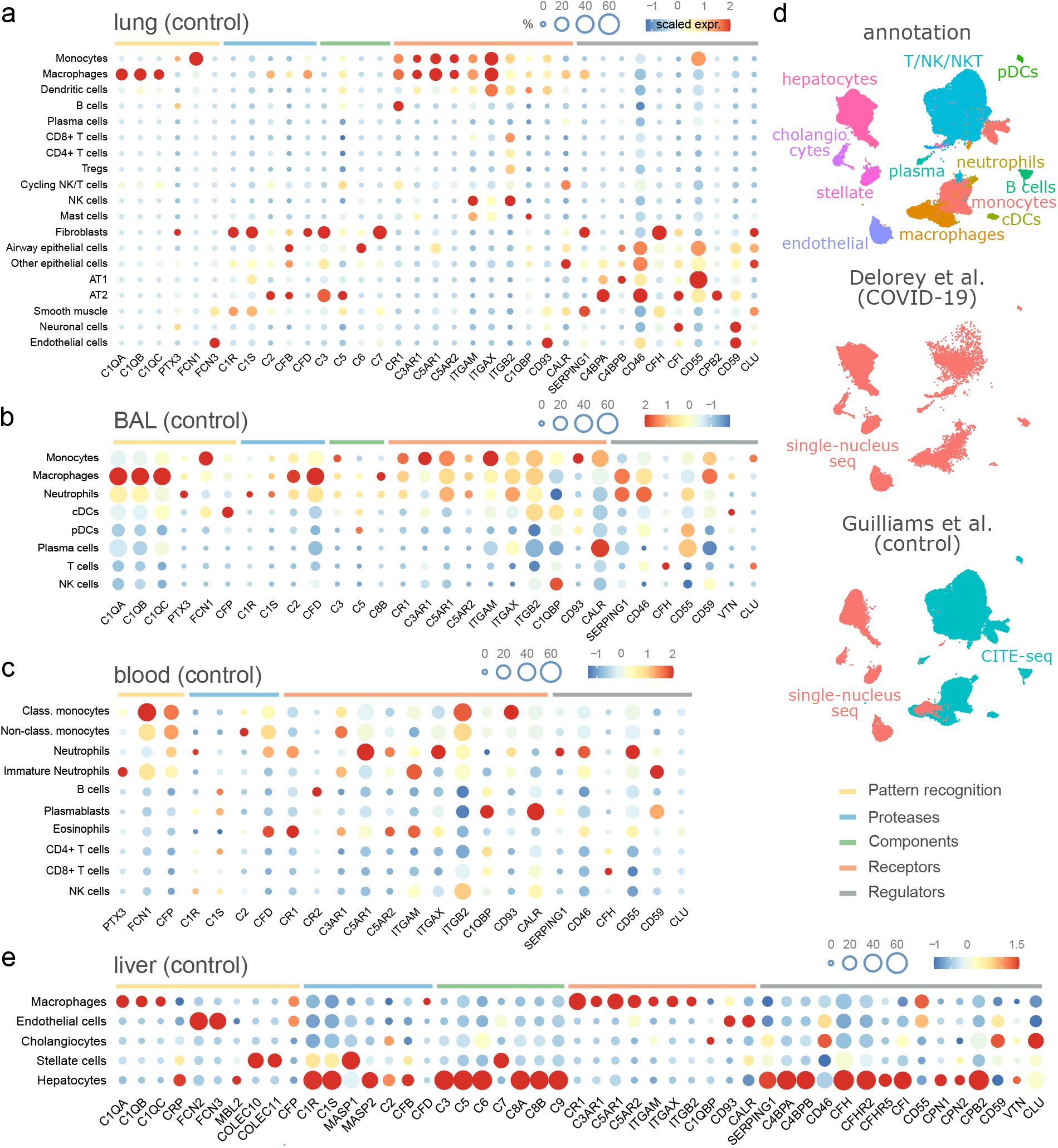
An atlas of complement gene expression across tissues and cell types in control samples. a, b, c, Expression of complement genes across control samples in lung (panel a), broncho-alveolar lavage (BAL) (panel b), fresh whole blood and peripheral-blood mononuclear cell (panel c). The dot size represents the proportion of cells expressing complement genes, while the color represents the average expression level. d, The Harmony-integrated liver dataset with both COVID-19 liver autopsies^45^ and control (healthy or obese)^46^ liver cells. Cell type annotation is displayed on top, single-nucleus sequencing data from COVID-19 patients in the middle, and control samples with both single-nucleus and CITE-sequencing data on the bottom. e, Complement gene expression in single-nucleus sequenced liver samples of non-COVID-19 patients in the merged dataset.

**Supplementary Figure 4:**
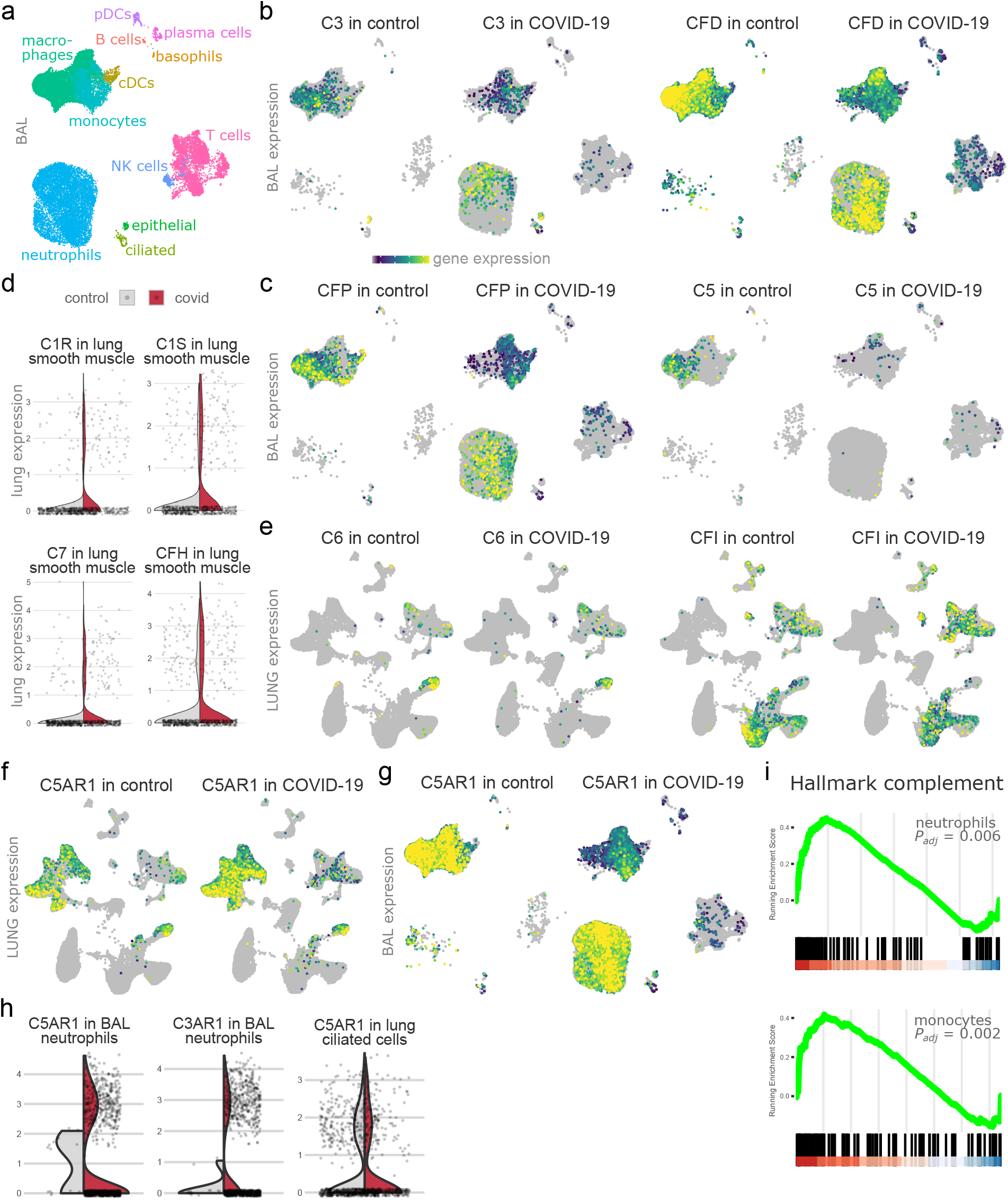
Complement production is divided between lung stroma, epithelium, myeloid cells and liver cells. a, Uniform manifold approximation and projection (UMAP) of CITE-sequencing of broncho-alveolar lavage (BAL) cells, which are colored by cell type annotation. b, c, g, Gene expression in BAL of C3, complement factor D (CFD) (b), complement factor properdin (CFP), C5 (c), and C5AR1 (g). d, h, Expression of complement factors (d) and receptors (h) in lung or BAL, represented as violin plots and split up for control and infected samples. A representative number of cells was overlaid. e, f, C6, CFI (e) and C5AR1 (f) expression plotted on the lung single-nucleus sequencing data. i, Gene set enrichment analysis in COVID-19 versus control patients of the Hallmark Complement gene set (Molecular Signatures Database). The upper plot depicts enrichment analysis BAL neutrophils, and the lower plot depicts monocytes in BAL.

**Supplementary Figure 5:**
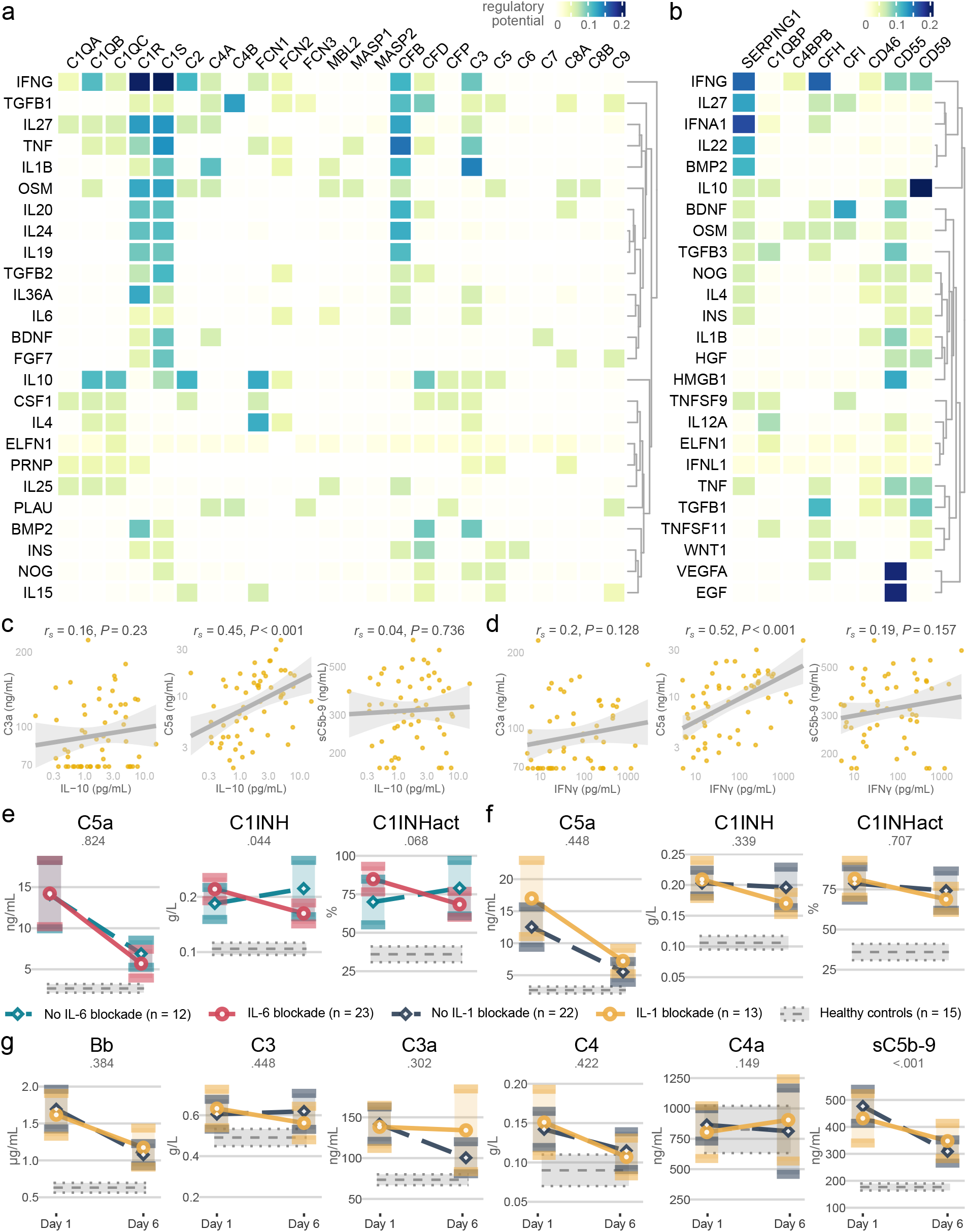
Interleukin 6 coordinates complement in COVID-19. a, b, Top 25 ligands with the highest predicted regulatory potential on the expression of complement factors (a) or complement regulators (b), according to the NicheNet ligand-target matrix^53^. c, d, Interleukin 10 (IL-10) (c) or interferon gamma (IFNγ) (d) serum levels in correlation to the activated complement components C3a, C5a and sC5b-9. The correlation coefficients and P values are calculated by two-sided Spearman’s rank tests. e, f, g, The evolution of complement components in COVID-19 patients who received anti-interleukin 6 (receptor) (e) or anti-interleukin 1 (f, g) versus control patients (in the COV-AID trial). Due to the factorial design, both groups contain patients treated with anti-IL-1 (e) or anti-IL-6(R) (f, g). The P values represent Wilcoxon testing performed on the change between day 1 and 6 of anti-IL-1 treated versus untreated participants. C1INHact: functional activity of C1 inhibitor.

**Supplementary Figure 6:**
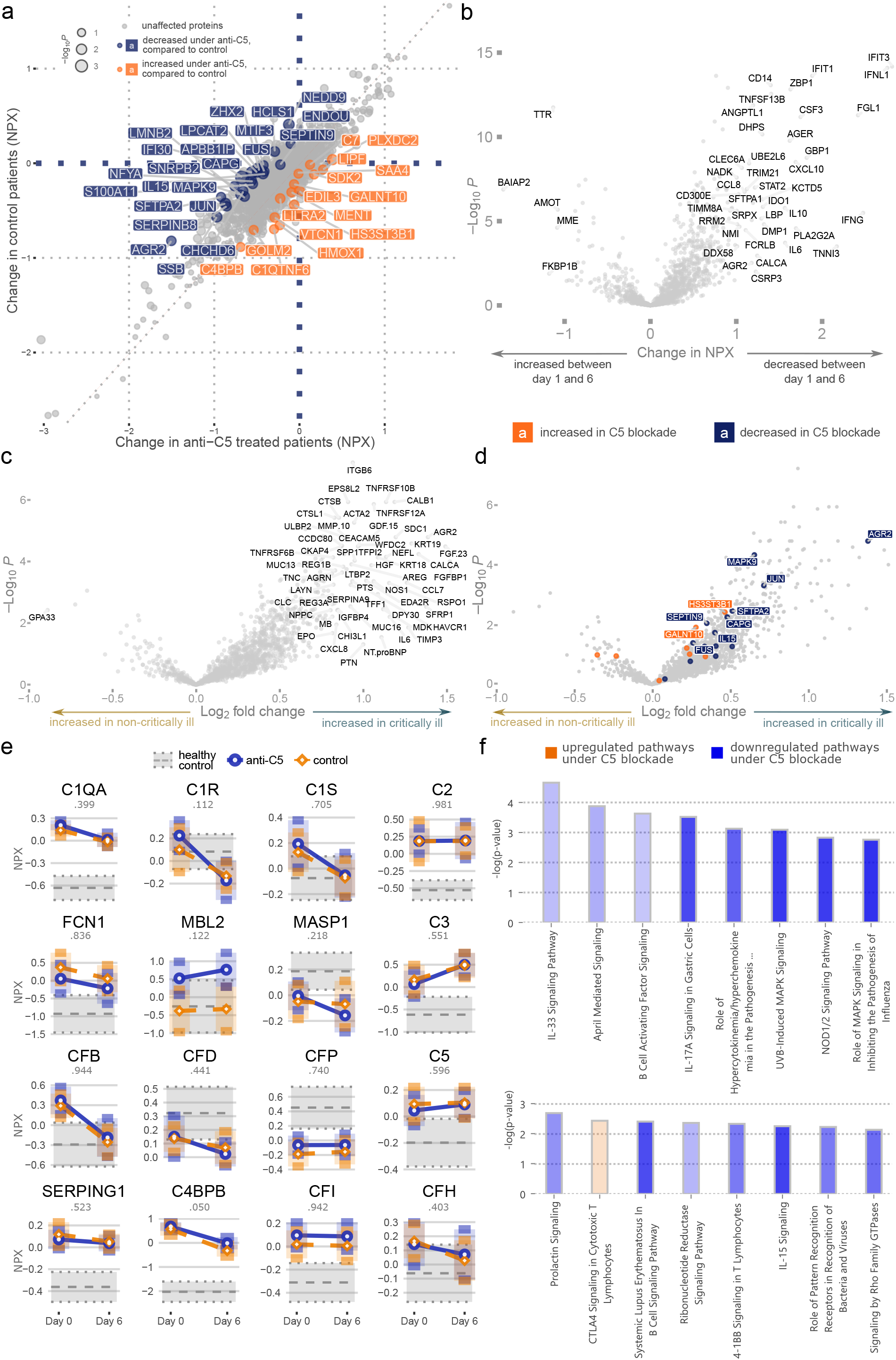
Complement inhibition improves the COVID-19 proteomic severity signature. a, Changes in serum proteins between day 1 and 6 in anti-C5 versus control patients, expressed as change in Normalized Protein eXpression (NPX). Proteins with a differential evolution, as determined by t-test P values < 0.05 (without adjustment for multiple testing) are plotted in blue (relative decrease in anti-C5 treated patients) or orange (relative increase under C5 inhibition). b, Evolution of proteins between day 1 and 6 in all COVID-19 patients. c, Markers associated with severe disease (WHO ordinal clinical severity scale of 4-5, oxygen therapy without invasive mechanical ventilation) versus critical illness (WHO ordinal clinical severity scale of 6-8, mechanical ventilation with or without additional support or death), from Byeon et al.^56^. The P values for b and c reflect by Benjamini-Hochberg corrected two-sided t-tests. d, Proteins altered by anti-C5 therapy (from Fig. 6b), overlaid on the independent critical illness-associated volcano plot from Byeon et al. (panel c). Blue proteins decrease in recipients of anti-C5 therapy, relative to untreated patients, while proteins in orange have a relative increase under C5 blockade. e, Changes over time in complement components under anti-C5 or control, in addition to healthy controls. P values reflect two-sided t-tests on the change between day 1 and 6 in control versus anti-C5 treated patients. f, Ingenuity Pathway Analysis of the protein changes in anti-C5 versus untreated COVID-19 patients. Fischer’s exact testing was used to generate P values.

Supplementary Table 1: Randomized clinical trial methods.

Supplementary Table 2: Patient characteristics of the complement level measurement.

Supplementary Table 3: Patient characteristics of the complement activity assays.

Supplementary Table 4: Patient characteristics of the Olink proteomic platform.

## Methods

### Trial participants, samples and clinical data

We conducted three randomized, open-label trials to test the efficacy and safety of various interventions in hospitalized adult COVID-19 patients across multiple hospitals in Belgium in 2020. In SARPAC^35^, we tested recombinant human granulocyte-macrophage colony-stimulating factor (twice daily rhu-GM-CSF, sargramostim via inhalation) to restore alveolar macrophage homeostasis in 81 COVID-19 patients with hypoxia and moderate inflammation (NCT04326920). In COV-AID^36^, we tested anti-interleukin drugs in 342 patients with hypoxia and severe inflammation due to COVID-19 (NCT04330638). The trial had a 2x2 factorial design, patients were sequentially randomized to IL-1 blockade (once daily recombinant interleukin 1 receptor antagonist, anakinra 100 mg subcutaneously) or no IL-1 blockade, and to IL-6 blockade (a single intravenous dose of either anti-IL- 6 receptor, tocilizumab 8 mg/kg or anti-IL-6, siltuximab 11 mg/kg) or no IL-6 blockade. In ZILUCOV, we studied the effect of C5 inhibition (daily zilucoplan 32.4 mg subcutaneously) or control in 81 patients with hypoxia and severe inflammation (NCT04382755). All patients in ZILUCOV received antibiotic prophylaxis against Neisseria meningitidis.

The full details on the in- and exclusion criteria, interventions and clinical outcomes of all three trials can be found in the published manuscripts and their trial protocols and statistical analysis plans^34–36^. In brief, all trials recruited patients with hypoxia (partial pressure of arterial oxygen divided by the fraction of inspired oxygen or PF ratio less than 350) and a recently confirmed COVID-19 infection. Enrollment in SARPAC was precluded if the ferritin level was above 2000 µg/L. COV-AID and ZILUCOV inclusion required high inflammatory markers (a) single ferritin measurement > 2000 μg/L on high flow oxygen or mechanical ventilation; (b) a ferritin concentration of > 1000 μg/L which had been increasing over the previous 24 h; (c) lymphopenia < 800/mL with two of the following: an increasing ferritin > 700 μg/L, an increasing lactate dehydrogenase > 300 IU/L, an increasing CRP > 70 mg/L, or an increasing D-dimers > 1000 ng/mL. (d) If the patient had three of the previous criteria at hospital admission with lymphopenia of less than 800/μL, there was no need to document an increase over 24 h. Immunosuppression for COVID-19 unrelated disorders or active co-infections were not permitted in COV-AID or ZILUCOV.

All trials were approved by the competent authorities, the appropriate Ethical Committees and conducted in accordance with Good Clinical Practice guidelines and the Declaration of Helsinki. All study subjects or their legal representatives provided oral and written consent for trial participation. Serum, plasma and peripheral blood mononuclear cell samples were collected in all trial participants on day 1 (before the first study dose, if applicable), on day 6 (before the 6^th^ dose of study, if applicable) and day 15 or at hospital discharge (whichever came first). Randomization, clinical data collection and data cleaning were carried out in the webbased system REDCap.

### Sample handling

Peripheral venous blood specimens were collected from healthy individuals and patients using simultaneously obtained EDTA and serum tubes. EDTA blood was diluted 1:2 in HBSS (Thermo Fisher Scientific; 24020117) and PBMCs isolated after gradient centrifugation over Ficoll-Paque (GE Healthcare; 17–1440-02). Cell-free plasma was subsequently transferred from the supernatant, aliquoted, and stored at −80°C. After two washings in cold HBSS, the yielded layer of PBMCs was counted in a Neubauer plate with trypan blue exclusion of dead cells. PBMCs were aliquoted in 90% FCS (Sigma-Aldrich; F7524) and 10% DMSO (Sigma-Aldrich; D2650). Vials were placed in a −80°C freezer using controlled rate freezing in preparation for final storage at −150°C until further use. Serum tubes were spun during 12 minutes at 2000g at 4°C, and cell-free serum was subsequently aliquoted and stored at −80°C until analysis.

### Complement factor measurement

Complement components Bb, C3a, C4a, C5a, and sC5b-9 were measured in cell-free plasma of SARPAC and COVAID participants with a customizable enzyme immunoassay multiplex kit (MicroVue Complement Multiplex; Quidel; A905s) according to the manufacturer’s instructions. Data were acquired on a Q-View Imager LS, using the Q-View Software 3.11. sC5b-9 levels in cell-free plasma of ZILUCOV participants were assessed using the MicroVue sC5b-9 plus enzyme immunoassay kit (A029). Absorbance was measured with the Emax Precision Microplate Reader with SoftMax Pro GxPv7.0.3 software. The plasma concentrations of C3, C4, and C1-INH were quantified by an automated turbidimetric assay on the Optilite analyzer (The Binding Site Group Limited), and C1-INH functional activity was measured using the Berichrom C1-Inhibitor assay (Siemens).

### Biomarker quantification

Serum cytokines were quantified using single- or multiplex immunoassays by Meso Scale Discovery (MSD). We measured IFN-γ, IL-1β, IL-6, IL-8, IL-10, IL-12p70, TNF-α using the V-PLEX Proinflammatory Panel 1 kit (1:2 dilution; K15049D), IP-10/CXCL10 by V-plex Chemokine Panel 1 (1:10; K151NVD), IL- 1RA by V-plex Cytokine Panel 2 (1:10; K151WTD), ICAM-1 by Vascular Injury Panel 2 (1:1000; K151SUD), GM-CSF by S-plex (1:2; K151F35), and G-CSF (B21VG) and IL-18 (B21VJ) using U-plex (1:2; K15227N). Data were acquired on a MESO QuickPlex SQ 120. Commercially available ELISA kits were used for the detection of serum Ang-2 (Thermo Fisher Scientific; KHC1641), sRAGE (R&D Systems; DRG00), MUC-1 (Thermo Fisher Scientific; EHMUC1), vWF (Thermo Fisher Scientific; EHVWF), suPAR (suPARnostic Virogates; E001). Samples were diluted at 1:5 (sRAGE), 1:10 (Ang-2 and vWF), 1:50 (MUC-1), 1:8000 (vWF) and analyzed according to the manufacturer’s instructions.

### SARS-CoV-2-specific antibody measurements

We quantified serum antibody levels directed against the SARS-CoV-2 spike 1 (S1) IgA (Euroimmun 2606-9601 A) and IgG (Euroimmun 2606-9601 G) and against the SARS-CoV-2 nucleocapsid (NCP) IgG (Euroimmun 2606-9601-2 G), according to the manufacturer’s protocol. Results were plotted as relative optical density versus the assay calibrator.

### Functional complement assay

The functional complement activity was measured on serum samples using the Wieslab Complement system Screen kit (Weislab Diagnostic Services; COMPL300 RUO), according to manufacturer’s protocol. Samples were diluted at 1:101, 1:101, and 1:18 for classical, lectin and alternative complement pathway, respectively.

### Sample size calculation and participant selection for proximity extension assay

The sample size calculation for proteomics was based on the preliminary observation of a 45% reduction in IL-8 in the anti-C5 treated group^34^. 20 replicates were required to detect a difference of 0.60 (equivalent to a reduction of 45%) at a significance level of 0.05, with a power of 0.844 using an F test. To select these samples, we first identified trial participants of the ZILUCOV with available unthawed serum samples from day 1 and day 6 (or discharge, whatever came first). We excluded patients with major protocol violations, such as missed doses within the first 5 days, or the absence of arterial blood gas sampling on day 1 or day 6. This yielded a total of 19 patients (out of 24) in the control arm and 49 patients (out of 54) in the anti-C5 arm. Although the randomization in clinical trial prevented selection bias, we carried out propensity score matching to achieve maximal baseline balance between control and intervention arm^72^. The propensity score matching was carried out using the R MatchIt v4.3.4 and optmatch v0.10.0 package. Optimal pair matching was used with a ratio of 1.1 and without replacement. The covariates for matching were age, gender, ethnicity, body mass index, history of diabetes mellitus, baseline ordinal scale, baseline SOFA score and baseline PaO_2_/FiO_2_. This led to adequately balanced arms in terms of propensity scores and included covariates. The full flowchart and participant characteristics are shown in Suppl. Fig. 2d and Supplementary Table 2.

### Proximity extension assay

Serum proteins were measured in the selected trial participants using the Olink Explore 3072 platform. This contained eight 384-plex panels: inflammation I&II, oncology I&II, cardiometabolic I&II and neurology I&II with panel lot numbers: B14806, B14807, B14808, B14809, B20704, B20705, B20706, B20707. Serum samples without previous thaw-freeze cycles of 8 healthy controls, 40 day 1 and 40 day 6 samples of ZILUCOV trial participants were plated in a randomized manner, refrozen and shipped to the Erasmus MC Olink Core Facility (Rotterdam, the Netherlands) for analysis. In the proximity extension assay, samples were incubated overnight with matched pairs of antibodies with unique DNA oligonucleotides. Hybridized oligonucleotide tails, brought in proximity due to target binding of both antibodies, were extended using a DNA polymerase. This was followed by a polymerase chain reaction (PCR), the addition of sample indexes and a second PCR step. Pooled DNA barcodes were purified and analyzed for quality control. Sequencing of the amplicons was performed with a NovaSeq 6000 system.

Data quantification was performed according to the platform guidelines. Sequence counts were quantified and normalized, yielding a Normalized Protein eXpression (NPX) unit, which is on a log2 scale. In total, 5 proteins were excluded for further analysis because of quality control reasons (SMAD1, ARHGAP25, KNG1, TNFSF9, TOM1L2). All other proteins were analyzed. Three proteins were normalized against the plate controls because of known bimodal distribution (TDGF1, FOLR3, PNLIPRP2). All other detected proteins (2921) were intensity normalized, as the plate-specific mean was subtracted from each sample on the respective plate. Specifically for the complement proteins, this method likely detects both the native as well as the activated proteins (personal communication by Olink).

### Proteomics and biomarker analysis

In general, all data wrangling and analysis was performed in R v4.2.1 using the tidyverse packages v1.3.2. The raw NPX results provided by Olink were extracted in a long format and integrated with the clinical and biomarker data. For proteins quantified multiple times (across multiple Olink panels), the average NPX was used for further analysis. The Olink data from Byeon et al.^56^ was downloaded from: https://www.thelancet.com/journals/landig/article/PIIS2589-7500(22)00112-1/fulltext.

To contrast proteomic results between two groups, we made volcano plots. We calculated average protein levels per group and subtracted these, yielding a log2(fold change) since Olink protein levels are expressed on a log2 scale. Visualizations were made with the EnhancedVolcano v1.14.0 and/or ggplot2 packages v3.4.0.

One complement sample was removed as outlier, with a sC5b-9 concentration of > 6.4 times the interquartile range (IQR) above the third quartile (Q3).

To plot protein or biomarker levels, we overlayed boxplots or violin plots with the individual datapoints in ggplot2. Evolutions over time were visualized as point-range plots, the 95% confidence intervals in these plots were calculated with the rstatix package v0.7.1. Scatterplots to investigate correlations were created in ggpubr v0.5.0. Heatmaps were made with the ComplexHeatmap package v2.12.1. Best correlations between single features and the whole dataframe were assessed using the lares package v5.1.4. Correlation matrices were created in the corrplot package v0.92. For visualization purposes, we used the packages viridis v0.6.2, ggthemes v4.2.4, ggrepel v0.9.3, scales v1.2.1 and RColorBrewer v1.1.3.

### Statistical testing

For the comparison of complement and cytokine levels or functional complement activities between three groups, we calculated Kruskal-Wallis rank tests with post-hoc pairwise Benjamini-Hochberg corrected Wilcoxon rank sum tests. To compare complement factor levels or activities between two groups, we calculated two-sided Wilcoxon tests. We computed Spearman’s rank correlation coefficients for correlations, and corrected these with the Benjamin-Hochberg method in the correlation matrices. Differences in Olink complement levels (expressed as NPX values) were evaluated with one-way ANOVA tests and follow-up Tukey’s honest significance difference tests. The evolution of complement factors (difference between day 1 and 6) under IL-6/IL-1 inhibition versus control were compared using two-sided Wilcoxon rank sum tests. All used statistical tests were two-sided and all plotted estimate ranges represent 95% confidence levels.

To focus the Olink proteomic analysis on COVID-19 relevant proteins, we prefiltered on the proteins which differed between healthy controls and COVID-19 patients at day 1. Given the log2- transformation and in line with previous literature^55, 56^, we assumed a normal distribution of the log2- transformed proteomic data. Based on unadjusted t-test P values < 0.05, we excluded 1393 proteins and retained 1529 for further analysis (Fig. 6a-d, Suppl. Fig. 6a-b). For the comparison of Olink protein levels between control and anti-C5 groups, we determined two-sided student t-tests without adjustment for multiplicity. The Olink protein changes over time were calculated by subtracting the NPX values (which equals to a log_2_ of the fold change) and we compared these changes for each group with student t-tests, without adjustment for multiplicity. In the paired analysis, we compared which proteins decreased or increased in one arm only. To this end, we calculated Benjamini-Hochberg corrected student t-tests for day 1 versus day 6 protein levels in each group, and then compared which proteins evolved differentially in each group. All statistical tests were carried out with the R base stats package v4.2.1.

### Single-cell sequencing data and analysis

All single-cell analysis was performed in R using Seurat v4.3.0^73^ and tidyverse packages. The COVID- 19 lung single-nucleus RNA sequencing atlas by Melms et al.^43^ was downloaded from: https://singlecell.broadinstitute.org/single_cell/study/SCP1219/columbia-university-nyp-covid-19-lung-atlas.

The broncho-alveolar lavage cells came from our own CITE-sequencing dataset which we previously published in combination with the SARPAC trial^35^. These data are available online via: https://www.single-cell.be/covid19/browser. We removed the non-SARS-CoV-2 infected patients and grouped the interstitial lung disease (1) with the control (2) samples and used these as non-infected controls.

Fresh blood, containing both whole and peripheral blood mononuclear cells from Schulte-Schrepping et al.^44^ were accessed via: https://beta.fastgenomics.org/datasets/detail-dataset-ee4b1a0f339140ad82f861aea35076f1.

Cluster annotation for lung, broncho-alveolar lavage and blood were copied from their respective publications^35, 43, 44^.

Single-nucleus RNA sequencing of COVID-19 liver samples published by Delorey et al.^45^ were retrieved via: https://singlecell.broadinstitute.org/single_cell/study/SCP1213/covid-19-liver-autopsy-samples. We first removed nuclei assigned as doublets, cycling, dying or without differentially expressed genes. We then combined this dataset with the human samples from our liver atlas, which contained both CITE-seq and single-nucleus data, published as Guilliams et al.^46^ For dataset merging, the raw data of both experiments were combined and normalized, the 2000 most variable genes were scaled and 40 principle components were calculated. To integrate the COVID-19 and control datasets, we ran Harmony according to the default settings with a maximum of 20 rounds of clustering^71^. Based on the Elbow Plot, we tested several numbers of principle components and created UMAPs based on 35 principle components. We clustered at a resolution of 0.8 and performed the further analysis after down sampling to 47000 cells per condition, to speed up the analysis. We manually reannotated the clusters and removed one additional cluster of doublets.

Gene expression plots per cell type were made using the Seurat DotPlot function, and further customized with ggplot2 v3.4.0. Only complement genes with expression in at least 0.5% of the cells were shown. For liver, we only plotted single-nucleus sequencing data. To visualize relative cell proportions in the stacked bar charts, we first downsampled the datasets to ensure an identical total cell number between the COVID-19 and control groups. We then plotted the relative contribution of each individual sample to the total number of cells per cell type. The split violin plots were created by extending the ggplot2 GeomViolin function as described under https://stackoverflow.com/questions/35717353/split-violin-plot-with-ggplot2. To contrast the number of cells in control versus COVID-19, we first downsampled each dataset to an equal number of cells per condition. Then, we plotted a fixed number of cells regardless of condition, revealing relative differences in cell numbers between both groups.

### Trajectory analysis

The diffusion map was created using the scanpy.tl.diffmap function of the Python Scanpy v1.5.1 package. Slingshot was applied on the first three diffusion components together the annotated clusters using the slingshot v1.4.0 R package. The Monocyte cluster was used as the starting point of the trajectory, as previously described^35^. The control and interstitial lung disease samples were merged. The COVID-19 samples (8) were randomly downsampled to the same cell number as non-infected samples. Samples from patients with bacterial infection were removed for the trajectory analysis. Line plots were created by plotting the gene expression according to the DC coordinates (DC1 was used for Fig. 4k, DC2 was used for Fig. 4i and 4j). Cells with outlier DC values were omitted (for Fig. 4i DC2 ≥ -0.0025 & DC2 ≤ 0.0135; for Fig. 4j DC2 ≥ -0.0045 & DC2 ≤ 0.0100; for Fig. 4k DC1 ≥ - 0.0050 & DC1 ≤ 0.0125). The geom_smooth function of the ggplot2 R package was used to draw the trend line, according to the default parameters.

### Upstream ligand prediction and gene signature analysis

The identification of ligands which could affect the expression of genes of interest was based on the NicheNet ligand-target data frame (https://zenodo.org/record/7074291/files/ligand_target_matrix_nsga2r_final.rds). This prior model integrated multiple data sources to quantify the likelihood that a particular ligand affects the expression of a particular target gene. We selected ligands which had the highest regulatory potential on average across the genes of interest, and plotted these with their regulatory potential per gene.

To determine the downstream signature induced by ligands of interest, NicheNet ligand activity analysis was performed. In this analysis, ligands are ranked according to how strongly genes with high regulatory potential of ligands are enriched in a gene set of interest compared to a background. The gene set of interest was determined as the upregulated genes in COVID-19 versus healthy controls for each cell type of interest; the background as all genes in NicheNet’s ligand-target matrix. To determine upregulated genes, muscat^74^, a state-of-the-art tool for differential state analysis in multi-group/multi-sample datasets, was used using the following thresholds: P value ≤ 0.05 and logFC ≥ 1. To compare ligand activity analyses between cell types, ligand activity values were z-score scaled prior to visualizations. Ligand-target links were inferred and visualized as described in the NicheNet software tutorials.

### Gene set enrichment analysis and pathway analysis

We performed gene set enrichment using the R clusterProfiler package v4.4.4^75^. Genes in the hallmark gene sets^76^ were retrieved via the msigdbr package v7.5.1. Differential gene of COVID-19 versus non-infected samples for specific cell types was calculated using the Seurat FindMarkers function.

QIAGEN Ingenuity Pathway Analysis (v84978992) was used to infer up- or downregulated pathways upon C5 blockade versus no C5 blockade. Proteins with an absolute log2 fold change of more than 0.25 were imported, with the user dataset (2894 mapped proteins, without prefiltering) as the reference set. Pathway Z-scores and P values (Fisher’s exact tests) were reported as computed by the software.

## Acknowledgements

We owe gratitude to all patients and their families for participating in our trials. We thank the VIB Tech Watch Fund and the VIB Grand Challenges Program, which enabled the proteomic and biomarker analysis. The respiratory diseases and primary immunodeficiency teams (Eva Van Braeckel, Pieter Depuydt, Stefanie Vermeersch, Benedicte Demeyere, Anja Delporte, Ans Vandecauter, Vanessa Parrein, Daisy Vermeiren, Karlien Claes) and the Health, innovation and research institute (HIRUZ) (Catherine Van Der Straeten, Charlotte Clauwaert, Joke Tommelein, Hélène De Naeyer, Dries Loncke, Kokur Hanife, Lieselot Vanlanduyt, Evy Doolaege, Simon Vanderschaeghe, Leen Borgers, Stefanie De Buyser) provided trial oversight and coordinated data collection, for which we thank them. Clinical samples were processed by many fantastic colleagues (Leen Sys, Helena Aegerter, Ursula Smole, Kim Deswarte, Leslie Naesens, Helena Flipts, Hamida Hammad, Veronique Debacker, Justine Van Moorleghem, Lisa Roels, Nancy Cabooter, Zara Declercq, Roanne Schuppers). We thank Claire Brittain, Laurent Detalle, Jemma Greenin and Marianna Lalla at UCB for the collaboration on the ZILUCOV trial. The Inflammation Research Center web team (Arne Soete, Kevin Verstaen) helped launching the Complement Atlas web tool. We thank Hamideh Baggali for providing linguistic advice on the manuscript.

## Author Contributions

KVD, LH, SJT and BNL conceptualized the study. KVD, LH, NV, PS and SJT performed experiments and analyzed data. KVD, JD, EDL, BM, CB and BNL coordinated the clinical trials and included study participants. KVD, LM, RB and SV analyzed the transcriptomic data. RC and STTS contributed to propensity matching and statistical analysis. STTS, MG, FH, SJT and BNL provided scientific guidance. KVD, LH and SJT drafted the first manuscript. All authors provided feedback and all approved the final publication.

## Data availability

All data produced in the present study are available upon reasonable request to the authors. Ethical statement

All studies were approved by the competent authorities and the Ethical Committee of Ghent University Hospital, and all trials were conducted in accordance with Good Clinical Practice guidelines and the Declaration of Helsinki.

## Additional Information

Supplementary Information is available for this paper.

## References

1. Wu, F. et al. A new coronavirus associated with human respiratory disease in China. Nature 579, 265–269 (2020).

2. Merad, M., Blish, C. A., Sallusto, F. & Iwasaki, A. The immunology and immunopathology of COVID-19. Science 375, 1122–1127 (2022).

3. Horby, P. et al. Dexamethasone in Hospitalized Patients with Covid-19. N. Engl. J. Med. 384, 693– 704 (2021).

4. Shankar-Hari, M. et al. Association Between Administration of IL-6 Antagonists and Mortality Among Patients Hospitalized for COVID-19. JAMA 326, 1–20 (2021).

5. Guimarães, P. O. et al. Tofacitinib in Patients Hospitalized with Covid-19 Pneumonia. N. Engl. J. Med. 385, 406–415 (2021).

6. Russell, C. D., Lone, N. I. & Baillie, J. K. Comorbidities, multimorbidity and COVID-19. Nat. Med. 29, 334–343 (2023).

7. Afzali, B., Noris, M., Lambrecht, B. N. & Kemper, C. The state of complement in COVID-19. Nat. Rev. Immunol. 22, 77–84 (2022).

8. Reis, E. S., Mastellos, D. C., Hajishengallis, G. & Lambris, J. D. New insights into the immune functions of complement. Nat. Rev. Immunol. 19, 503–516 (2019).

9. Ram, S., Lewis, L. A. & Rice, P. A. Infections of People with Complement Deficiencies and Patients Who Have Undergone Splenectomy. Clin. Microbiol. Rev. 23, 740–780 (2010).

10. Naesens, L. et al. Plasma C3d levels as a diagnostic marker for complete complement factor I deficiency. J. Allergy Clin. Immunol. 147, 749–753.e2 (2021).

11. Ma, L. et al. Increased complement activation is a distinctive feature of severe SARS-CoV-2 infection. Sci. Immunol. 6, 1–18 (2021).

12. Ramlall, V. et al. Immune complement and coagulation dysfunction in adverse outcomes of SARS-CoV-2 infection. Nat. Med. 26, 1609–1615 (2020).

13. Skendros, P. et al. Complement and tissue factor–enriched neutrophil extracellular traps are key drivers in COVID-19 immunothrombosis. J. Clin. Invest. 130, 6151–6157 (2020).

14. Carvelli, J. et al. Association of COVID-19 inflammation with activation of the C5a–C5aR1 axis. Nature 588, 146–150 (2020).

15. Hultström, M. et al. Genetic determinants of mannose-binding lectin activity predispose to thromboembolic complications in critical COVID-19. Nat. Immunol. 23, 861–864 (2022).

16. Holter, J. C. et al. Systemic complement activation is associated with respiratory failure in COVID- 19 hospitalized patients. Proc. Natl. Acad. Sci. 117, 25018–25025 (2020).

17. Stravalaci, M. et al. Recognition and inhibition of SARS-CoV-2 by humoral innate immunity pattern recognition molecules. Nat. Immunol. 23, 275–286 (2022).

18. Castanha, P. M. S. et al. IgG response to SARS-CoV-2 and seasonal coronaviruses contributes to complement overactivation in severe COVID-19 patients. J. Infect. Dis. 54, 1–54 (2022).

19. Devalaraja-Narashimha, K. et al. Association of complement pathways with COVID-19 severity and outcomes. Microbes Infect. 105081 (2022) doi:10.1016/j.micinf.2022.105081.

20. Siggins, M. K. et al. Alternative pathway dysregulation in tissues drives sustained complement activation and predicts outcome across the disease course in COVID-19. Immunology 168, 473– 492 (2022).

21. Boussier, J. et al. Severe COVID-19 is associated with hyperactivation of the alternative complement pathway. J. Allergy Clin. Immunol. 149, 550–556.e2 (2022).

22. Satyam, A. et al. Activation of classical and alternative complement pathways in the pathogenesis of lung injury in COVID-19. Clin. Immunol. 226, 108716 (2021).

23. Gao, T. et al. Highly pathogenic coronavirus N protein aggravates inflammation by MASP-2- mediated lectin complement pathway overactivation. Signal Transduct. Target. Ther. 7, 1–15 (2022).

24. Ali, Y. M. et al. Lectin Pathway Mediates Complement Activation by SARS-CoV-2 Proteins. Front. Immunol. 12, 1–8 (2021).

25. Liszewski, M. K. et al. Intracellular Complement Activation Sustains T Cell Homeostasis and Mediates Effector Differentiation. Immunity 39, 1143–1157 (2013).

26. Georg, P. et al. Complement activation induces excessive T cell cytotoxicity in severe COVID-19. Cell 185, 493–512.e25 (2022).

27. Chaudhary, N., Jayaraman, A., Reinhardt, C., Campbell, J. D. & Bosmann, M. A single-cell lung atlas of complement genes identifies the mesothelium and epithelium as prominent sources of extrahepatic complement proteins. Mucosal Immunol. 15, 927–939 (2022).

28. Sahu, S. K. et al. Lung epithelial cell–derived C3 protects against pneumonia-induced lung injury. Sci. Immunol. 8, eabp9547 (2023).

29. Tam, J. C. H., Bidgood, S. R., McEwan, W. A. & James, L. C. Intracellular sensing of complement C3 activates cell autonomous immunity. Science 345, 1256070 (2014).

30. Kulkarni, H. S. et al. Intracellular C3 Protects Human Airway Epithelial Cells from Stress- associated Cell Death. Am. J. Respir. Cell Mol. Biol. 60, 144–157 (2019).

31. Vlaar, A. P. J. et al. Anti-C5a antibody (vilobelimab) therapy for critically ill, invasively mechanically ventilated patients with COVID-19 (PANAMO): a multicentre, double-blind, randomised, placebo-controlled, phase 3 trial. Lancet Respir. Med. 10, 1137–1146 (2022).

32. Skendros, P. et al. Complement C3 inhibition in severe COVID-19 using compstatin AMY-101. Sci. Adv. 8, eabo2341 (2022).

33. Carvelli, J. et al. Avdoralimab (Anti-C5aR1 mAb) Versus Placebo in Patients With Severe COVID- 19: Results From a Randomized Controlled Trial (FOR COVID Elimination [FORCE]). Crit. Care Med. 50, 1788–1798.

34. De Leeuw, E. et al. Efficacy and safety of the investigational complement C5 inhibitor zilucoplan in patients hospitalized with COVID-19: an open-label randomized controlled trial. Respir. Res. 23, 202 (2022).

35. Bosteels, C. et al. Loss of GM-CSF-dependent instruction of alveolar macrophages in COVID-19 provides a rationale for inhaled GM-CSF treatment. Cell Rep. Med. 3, 100833 (2022).

36. Declercq, J. et al. Effect of anti-interleukin drugs in patients with COVID-19 and signs of cytokine release syndrome (COV-AID): a factorial, randomised, controlled trial. Lancet Respir. Med. 9, 1427–1438 (2021).

37. Giamarellos-Bourboulis, E. J. et al. Complex Immune Dysregulation in COVID-19 Patients with Severe Respiratory Failure. Cell Host Microbe 27, 992–1000.e3 (2020).

38. Al-Nesf, M. A. Y. et al. Prognostic tools and candidate drugs based on plasma proteomics of patients with severe COVID-19 complications. Nat. Commun. 13, 1–14 (2022).

39. Desvaux, E. et al. Network-based repurposing identifies anti-alarmins as drug candidates to control severe lung inflammation in COVID-19. PLOS ONE 16, e0254374 (2021).

40. Yan, B. et al. SARS-CoV-2 drives JAK1/2-dependent local complement hyperactivation. Sci. Immunol. 6, 1–15 (2021).

41. Arbore, G. et al. T helper 1 immunity requires complement-driven NLRP3 inflammasome activity in CD4+ T cells. Science 352, aad1210 (2016).

42. Niyonzima, N., et al. Mitochondrial C5aR1 activity in macrophages controls IL-1β production underlying sterile inflammation. Sci. Immunol. 6, eabf2489 (2021).

43. Melms, J. C. et al. A molecular single-cell lung atlas of lethal COVID-19. Nature 595, 114–119 (2021).

44. Schulte-Schrepping, J. et al. Severe COVID-19 Is Marked by a Dysregulated Myeloid Cell Compartment. Cell 182, 1419–1440.e23 (2020).

45. Delorey, T. M. et al. COVID-19 tissue atlases reveal SARS-CoV-2 pathology and cellular targets. Nature 595, 107–113 (2021).

46. Guilliams, M. et al. Spatial proteogenomics reveals distinct and evolutionarily conserved hepatic macrophage niches. Cell 185, 379–396.e38 (2022).

47. Sefik, E. et al. Inflammasome activation in infected macrophages drives COVID-19 pathology. Nature 606, 585–593 (2022).

48. Chen, S. T. et al. A shift in lung macrophage composition is associated with COVID-19 severity and recovery. Sci. Transl. Med. 14, eabn5168 (2022).

49. The NU SCRIPT Study Investigators, et al. Circuits between infected macrophages and T cells in SARS-CoV-2 pneumonia. Nature 590, 635–641 (2021).

50. Street, K. et al. Slingshot: cell lineage and pseudotime inference for single-cell transcriptomics. BMC Genomics 19, 477 (2018).

51. Aegerter, H., Lambrecht, B. N. & Jakubzick, C. V. Biology of lung macrophages in health and disease. Immunity 55, 1564–1580 (2022).

52. PA Kager, CE Hack, AJ Hannema, & PH Rees. High Clq Levels, Low Cls/Clq Ratios, and High Levels of Circulating Immune Complexes in Kala Azar. Clin. Immunol. Immunopathol. 23, 86–93 (1982).

53. Browaeys, R., Saelens, W. & Saeys, Y. NicheNet: modeling intercellular communication by linking ligands to target genes. Nat. Methods 17, 159–162 (2020).

54. Lim, E. H. T. et al. Anti-C5a antibody vilobelimab treatment and the effect on biomarkers of inflammation and coagulation in patients with severe COVID-19: a substudy of the phase 2 PANAMO trial. Respir. Res. 23, 375 (2022).

55. Filbin, M. R. et al. Longitudinal proteomic analysis of severe COVID-19 reveals survival-associated signatures, tissue-specific cell death, and cell-cell interactions. Cell Rep. Med. 2, 100287 (2021).

56. Byeon, S. K. et al. Development of a multiomics model for identification of predictive biomarkers for COVID-19 severity: a retrospective cohort study. Lancet Digit. Health 4, e632–e645 (2022).

57. Sinha, P., Meyer, N. J. & Calfee, C. S. Biological Phenotyping in Sepsis and Acute Respiratory Distress Syndrome. Annu. Rev. Med. 74, 457–471 (2023).

58. van der Zee, P., Rietdijk, W., Somhorst, P., Endeman, H. & Gommers, D. A systematic review of biomarkers multivariately associated with acute respiratory distress syndrome development and mortality. Crit. Care 24, 243 (2020).

59. Beitler, J. R. et al. Advancing precision medicine for acute respiratory distress syndrome. Lancet Respir. Med. 10, 107–120 (2022).

60. Standiford, T. J. & Ward, P. A. Therapeutic targeting of acute lung injury and acute respiratory distress syndrome. Transl. Res. 167, 183–191 (2016).

61. Bosmann, M. et al. Extracellular histones are essential effectors of C5aR- and C5L2-mediated tissue damage and inflammation in acute lung injury. FASEB J. 27, 5010–5021 (2013).

62. Halpern, K. B. et al. Single-cell spatial reconstruction reveals global division of labour in the mammalian liver. Nature 542, 352–356 (2017).

63. Kee, J. et al. SARS-CoV-2 disrupts host epigenetic regulation via histone mimicry. Nature 610, 381–388 (2022).

64. Kumar, J., Dhyani, S., Kumar, P., Sharma, N. R. & Ganguly, S. SARS-CoV-2-encoded ORF8 protein possesses complement inhibitory properties. J. Biol. Chem. 299, 102930 (2023).

65. Paludan, S. R. & Mogensen, T. H. Innate immunological pathways in COVID-19 pathogenesis. Sci. Immunol. 7, eabm5505 (2022).

66. Yu, J. et al. Direct activation of the alternative complement pathway by SARS-CoV-2 spike proteins is blocked by factor D inhibition. Blood 136, 2080–2089 (2020).

67. Kilgore, K. S., Ward, P. A. & Warren, J. S. Neutrophil Adhesion to Human Endothelial Cells is Induced by the Membrane Attack Complex: The Roles of P-Selectin and Platelet Activating Factor. Inflammation 22, 583–598 (1998).

68. Mantovani, A. & Garlanda, C. Humoral Innate Immunity and Acute-Phase Proteins. N. Engl. J. Med. 388, 439–452 (2023).

69. Hack, C. E. et al. Elevated plasma levels of the anaphylatoxins C3a and C4a are associated with a fatal outcome in sepsis. Am. J. Med. 86, 20–26 (1989).

70. Palma Medina, L. M., et al. Targeted plasma proteomics reveals signatures discriminating COVID- 19 from sepsis with pneumonia. Respir. Res. 24, 62 (2023).

71. Korsunsky, I. et al. Fast, sensitive and accurate integration of single-cell data with Harmony. Nat. Methods 16, 1289–1296 (2019).

72. Ho, D. E., Imai, K., King, G. & Stuart, E. A. Matching as Nonparametric Preprocessing for Reducing Model Dependence in Parametric Causal Inference. Polit. Anal. 15, 199–236 (2007).

73. Hao, Y. et al. Integrated analysis of multimodal single-cell data. Cell 184, 3573–3587.e29 (2021).

74. Crowell, H. L. et al. muscat detects subpopulation-specific state transitions from multi-sample multi-condition single-cell transcriptomics data. Nat. Commun. 11, 6077 (2020).

75. Wu, T. et al. clusterProfiler 4.0: A universal enrichment tool for interpreting omics data. The Innovation 2, 100141 (2021).

76. Liberzon, A. et al. The Molecular Signatures Database (MSigDB) hallmark gene set collection. Cell Syst. 1, 417–425 (2015).

