## Supplementary Tables 1-4 for "A Complement Atlas identifies interleukin 6 dependent alternative pathway dysregulation as a key druggable feature of COVID-19"

**Supplementary Table 1**: Randomized clinical trial methods.

<table>
<tr><th></th><th>SARPAC</th><th>COV-AID</th><th>ZILUCOV</th></tr>
<tr><td>Full publication</td><td>https://doi.org/10.1016/j.xcrm.2022.100833</td><td>https://doi.org/10.1016/S2213-2600(21)00377-5</td><td>https://doi.org/10.1186/s12931-022-02126-2</td></tr>
<tr><td>Registration</td><td>NCT04326920<br>2020-001254-22</td><td>NCT04330638<br>2020-001500-4</td><td>NCT04382755<br>2020-00213033</td></tr>
<tr><td>Inclusion criteria</td><td></td><td></td><td></td></tr>
<tr><td>COVID-19</td><td>Proven SARS-CoV-2 infection ≤ 2 weeks</td><td colspan="2">Proven SARS-CoV-2 infection, symptoms between 6 and 16 days</td></tr>
<tr><td>Oxygenation</td><td>PF ratio < 350 mmHg or $SpO_2$ < 93% on 2L $O_2$<br>-</td><td>PF ratio < 350 mmHg<br>Bilateral pulmonary infiltrates</td><td>PF ratio < 350 mmHg or $SpO_2$ < 93% on 2L $O_2$<br>Bilateral pulmonary infiltrates</td></tr>
<tr><td>Inflammation</td><td>Ferritin < 2000 µg/L</td><td colspan="2">Ferritin > 2000 µg/L if on HFNO or IMV<br>OR Ferritin > 1000 µg/L and increasing over previous 24h<br>OR Lymphopenia < 800/µL with two of the following criteria: (1) increasing ferritin > 700 µg/L, (2) increasing LDH > 300 IU/L, (3) increasing CRP > 70 mg/L, (4) increasing D-dimers > 1000 ng/mL<br>OR If lymphopenia with 3 of the above were present at hospital admission, there was no need to document a rise</td></tr>
<tr><td>Age</td><td>Between 18 and 80 years</td><td colspan="2">> 18 years</td></tr>
<tr><td>Exclusion criteria</td><td></td><td></td><td></td></tr>
<tr><td>Ventilation</td><td>Mechanical ventilation at inclusion</td><td colspan="2">Mechanical ventilation > 24 h at randomisation, or ECMO at randomisation</td></tr>
<tr><td>Frailty</td><td>-</td><td colspan="2">Clinical frailty score > 3 before SARS-CoV-2 infection</td></tr>
<tr><td>Critical illness</td><td>-</td><td colspan="2">Unlikelihood of survival beyond 48 h</td></tr>
<tr><td>Co-infections</td><td>-</td><td colspan="2">Active co-infection</td></tr>
<tr><td>Hematologic</td><td>White blood cell count > 25 000/µL</td><td colspan="2">Trombocytopenia < 50 000/µL or neutropenia < 1500/µL</td></tr>
<tr><td>Medical history</td><td>Active myeloid malignancy</td><td>History of bowel perforation of diverticulitis</td><td>History of meningococcal disease</td></tr>
<tr><td>Immunosuppression</td><td>> 20 mg methylprednisolone or equivalent for a COVID-19 unrelated disorder</td><td colspan="2">High-dose systemic steroid (> 8 mg methylprednisolone or equivalent for > 1 month) or immunosuppressive drug for a COVID-19 unrelated disorder</td></tr>
<tr><td>Allergy</td><td colspan="3">History of serious allergic reactions to any of the study drugs</td></tr>
<tr><td>Other</td><td colspan="3">Pregnancy or breastfeeding</td></tr>
<tr><td>Trial design</td><td>Phase 2, multi-center, open-label</td><td>Phase 3, 2x2 factorial design, multi-center, open-label</td><td>Phase 2, multi-center, open-label</td></tr>
<tr><td>Timing</td><td>March 24, 2020 to September 8, 2020</td><td>April 4, 2020 to Dec 6, 2020</td><td>Aug 15, 2020 to Dec 16, 2020</td></tr>
<tr><td>Randomization</td><td>1:1 for GM-CSF or no GM-CSF<br>Via interactive Web Response System (REDCap)</td><td>1:2 for anakinra or no anti-IL-1<br>1:1:1 for tocilizumab, siltuximab or no anti-IL-6(R)<br>Via interactive Web Response System (REDCap)</td><td>2:1 for anti-C5 or no anti-C5<br>Via interactive Web Response System (REDCap)</td></tr>
<tr><td>Intervention</td><td>Twice daily sargramostim (125 µg) via inhalation</td><td>Once daily anakinra (100 mg) subcutaneously*</td><td></td></tr>
</table>

| | | | |
|---|---|---|---|
| | | Single dose of tocilizumab (8 mg/kg) intravenously<br>Single dose of siltuximab (11 mg/kg) intravenously | Once daily zilucoplan (32,4 mg) subcutaneously**<br>Prophylactic antibiotics covering N. menigitidis in all patients |
| Participants | 81 | 342 | 81 |

* For 28 days or until hospital discharge, whichever came first

** For 14 days or until hospital discharge, whichever came first

ECMO: extracorporeal membrane oxygenation; HFNO: high-flow nasal oxygen; IMV: invasive mechanical ventilation

**Supplementary Table 2**: Patient characteristics of the complement level measurement.

| *Characteristics of patients with day 1 data* | **SARPAC**<br>N = 59 | **COV-AID**<br>N = 35 | **Healthy**<br>N = 15 |
|---|---|---|---|
| Male sex - no. (%) | 35 (59.3) | 25 (71.4) | 9 (60.0) |
| Age - mean (SD), years | 61 (12.0) | 63.7 (12.8) | 57.6 (8.6) |
| Diabetes mellitus† - no. (%) | 13 (46.4) | 10 (29.4) | |
| 6-point ordinal scale at baseline - no. (%) | | | |
| 2 Invasive mechanical ventilation | 1 (1.7) | 4 (11.4) | |
| 3 Non-invasive ventilation or high flow oxygen devices | 5 (8.5) | 7 (20) | |
| 4 Hospitalized, requiring supplemental oxygen | 51 (86.4) | 24 (68.6) | |
| 5 Hospitalized, no supplemental oxygen | 2 (3.4) | - | |
| Laboratory values at baseline - mean (SD) | | | |
| CRP - mg/L | 102.7 (75.7) | 166.5 (96.4) | |
| Ferritin - µg/L | 684 (498.2) | 2138 (1224.9) | |
| D-dimers - ng/mL | 736.3 (346.6) | 1403 (1177.6) | |
| PaO2/FiO2 ratio at baseline - mean (SD), mmHg | 283.6 (61.7) | 202.1 (86.9) | |
| Trial intervention - no. (%) | GM-CSF - 31 (52.5)<br>No GM-CSF - 28 (47.5) | Control - 7 (20)<br>Anti-IL-1 - 5 (14.3)<br>Anti-IL6 or anti-IL6R - 15 (42.9)<br>Anti-IL-1 and anti-IL-6(R) - 8 (22.9) | |
| All-cause mortality‡ - no. (%) | 2 (3.4) | 2 (5.7) | |
| Need for invasive mechanical ventilation at any moment‡ - no. (%) | 5 (8.6) | 12 (34.3) | |
| Severity classification‡* - no (%) | | | |
| Severe | 53 (91.4) | 23 (65.7) | |
| Critical | 5 (8.6) | 12 (34.3) | |

† One missing in both the SARPAC and COV-AID cohorts

‡ One missing in SARPAC

* Critical disease was defined as the need for mechanical ventilation at any time or resulting in death

**Supplementary Table 3**: Patient characteristics of the complement activity assays.

| | SARPAC<br>N = 13 | COV-AID<br>N = 18 | ZILUCOV<br>N = 22 | Healthy<br>N = 10 |
|---|---|---|---|---|
| Male sex - no. (%) | 7 (54) | 15 (83.3) | 18 (81.8) | 9 (90) |
| Age - mean (SD), years | 58.9 (16) | 62.5 (10.1) | 64.3 (10.6) | 59.1 (7.1) |
| Diabetes mellitus - no. (%) | 2 (15.4) | 4 (22.2) | 4 (18.2) | |
| 6-point ordinal scale at baseline - no. (%) | | | | |
| 2 Invasive mechanical ventilation | 1 (7.7) | 1 (5.6) | 1 (1.5) | |
| 3 Non-invasive ventilation or high flow oxygen devices | 1 (7.7) | 6 (33.3) | 8 (36.4) | |
| 4 Hospitalized, requiring supplemental oxygen | 11 (84.6) | 11 (61.1) | 13 (59.1) | |
| PaO2/FiO2 ratio at baseline - mean (SD), mmHg | 258.2 (66.3) | 198.9 (89.9) | 171.8 (94.4) | |
| Glucocorticoid use (during first 5 days) - no. (%) | 13 (100) | 18 (100) | 20 (90.9) | |
| Trial intervention - no. (%) | Control arm - 13 (100) | Anti-IL6(R) in all Siltuximab - 8 (44.4) Tocilizumab - 10 (55.6) | Control arm - 22 (100) | |
| All-cause mortality - no. (%) | 0 (0) | 3 (16.7) | 5 (22.7) | |
| Time until 2-point improvement on 6-point ordinal scale or hospital discharge* - mean (range), days | NA | 15.5 (4, 28) | 16.8 (6, 28) | |
| Time until hospital discharge* - mean (range), days | 16 (6, 28) | 15.4 (5, 28) | 18,1 (6, 28) | |
| Need for invasive mechanical ventilation at any moment - no. (%) | 4 (30.7) | 7 (38.9) | 6 (27.3) | |
| Severity classification** - no (%) | | | | |
| Severe | 9 (69.2) | 11 (61.1) | 15 (68.2) | |
| Critical | 4 (30.7) | 7 (38.9) | 7 (31.8) | |

* Patients who died or did not recover within 28 days were censored at day 28

** Critical disease was defined as the need for mechanical ventilation at any time or resulting in death

NA = not available

**Supplementary Table 4**: Patient characteristics of the Olink proteomic platform.

| *Baseline characteristics* | **Zilucoplan** N = 21 | **Standard of Care** N = 19 | **Healthy** N = 8 |
|---|---|---|---|
| Male sex - no. (%) | 17 (81) | 16 (84.2) | 6 (80) |
| Ethnicity - no. (%) | | | |
| African | 1 (4.8) | - | - |
| Arabian | - | 1 (5.3) | - |
| Caucasian | 20 (95.2) | 17 (89.5) | 8 (100) |
| Other | - | 1 (5.3) | - |
| Age - mean (SD), years | 66.2 (11) | 65.1 (10.9) | 56.3 (7.2) |
| BMI - mean (SD) | 29.9 (3.8) | 30.9 (4.7) | 28.8 (1.9) |
| Co-existing conditions - no. (%) | | | |
| Arterial hypertension | 8 (38.1) | 8 (42.1) | |
| Diabetes mellitus | 4 (19) | 4 (21.1) | |
| Cardiovascular disease | 4 (19) | 9 (47.4) | |
| Chronic kidney disease | 2 (9.5) | 1 (5.3) | |
| 6-point ordinal scale at baseline - no. (%) | | | |
| 2 Invasive mechanical ventilation | 2 (9.5) | 1 (5.3) | |
| 3 Non-invasive ventilation or high flow oxygen devices | 6 (28.6) | 7 (36.8) | |
| 4 Hospitalized, requiring supplemental oxygen | 13 (61.9) | 11 (57.9) | |
| Admitted to ICU at randomisation - no. (%) | 11 (52.4) | 9 (47.4) | |
| Days of symptoms at randomisation - median (min, max) | 9 (6, 15) | 7.5 (5, 12) | |
| Days of hospitalization at randomisation - median (min, max) | 2 (0, 6) | 1 (0, 6) | |
| PaO2/FiO2 ratio at baseline - mean (SD), mmHg | 166.9 (88.9) | 172.7 (98.8) | |
| A-a gradient at baseline - mean (SD), mmHg | 243.2 (179.2) | 261.8 (219.9) | |
| SOFA score at baseline* - no. (%) | | | |
| 1-2 | 12 (63.2) | 11 (57.9) | |
| 3-4 | 4 (21.1) | 6 (31.6) | |
| 5-6 | 1 (5.3) | 2 (10.5) | |
| 7 | 2 (10.5) | - | |
| Laboratory values at baseline - mean (SD) | | | |
| CRP - mg/L | 126.7 (53.8) | 132.5 (59.2) | |
| Lymphocyte count - 10^9/L | 0.80 (0.4) | 0.64 (0.4) | |
| Ferritin - µg/L | 2023.2 (946.6) | 2023.9 (1014.4) | |

| | | |
|---|---|---|
| D-dimers** - ng/mL | 1029.3 (826.9) | 1132.6 (911.6) |
| LDH - IU/L | 422.9 (140.4) | 505.2 (179.1) |
| C5 - µg/L | 111.4 (18.4) | 94.7 (24.1) |
| Concomitant medication - no. (%) | | |
| Glucocorticoids (at randomisation) | 15 (71.4) | 10 (52.6) |
| Glucocorticoid use (during first 5 days) | 16 (76.2) | 14 (73.7) |
| Anticoagulants (at randomisation) | 13 (61.9) | 11 (57.9) |
| Remdesivir (at randomisation) | 2 (9.5) | 2 (10.5) |

* 2 baseline SOFA scores are missing in the zilucoplan arm

** One D-dimer value was excluded, with a concentration of 554 times the IQR above Q3

| *Outcomes* | **Zilucoplan** | | **Control** | |
|---|---|---|---|---|
| | No. available | | No. available | |
| All-cause mortality - no. (%) | 21 | 3 (14.3) | 19 | 5 (26.3) |
| Time until 2-point improvement on 6-point ordinal scale or hospital discharge - median (range), days | 16 | 15 (5, 23) | 12 | 11 (6, 21) |
| Need for invasive mechanical ventilation at any moment - no. (%) | 21 | 4 (19.0) | 19 | 6 (31.6) |
| Change from baseline in PaO2/FiO2 - mean (SD), mmHg | | | | |
| At day 6 | 21 | +51.4 (66.1) | 19 | +16.2 (118.9) |
| At day 15 | 19 | +126.9 (102.5) | 17 | +76.0 (133.3) |
| Change from baseline in A-a gradient - mean (SD), mmHg | | | | |
| At day 6 | 21 | -61.3 (138.2) | 19 | -33.3 (183.3) |
| At day 15 | 16 | -182.5 (168.7) | 17 | -103.9 (191.1) |
| Duration of hospital stay since randomization - average (SD), days | 18 | 17.1 (10.8) | 14 | 16.2 (10.5) |
| Severity classification* - no (%) | 21 | | 19 | |
| Severe | | 17 (81.0) | | 12 (63.1) |
| Critical | | 4 (19.0) | | 7 (36.8) |
| Distance walked in a 6-minute walk test at follow-up** - mean (SD) meters | 18 | 506.9 (102.7) | 13 | 485.1 (148.3) |
| WHO performance category at follow-up** - no (%) | 18 | | 13 | |
| 0 | | 10 (55.6) | | 7 (53.8) |
| 1 | | 8 (44.4) | | 5 (38.5) |
| ≥2 | | 0 | | 1 (7.7) |
| HRCT score at follow-up** - mean (SD) | 9 | 107.6 (6.8) | 10 | 111 (20.9) |

* Critical disease was defined as the need for mechanical ventilation at any time or resulting in death

** Follow-up visit occurred between 12 and 22 weeks post-randomization
